# Rates of serious clinical outcomes in survivors of hospitalisation with COVID-19: a descriptive cohort study within the OpenSAFELY platform

**DOI:** 10.1101/2021.01.22.21250304

**Authors:** The OpenSAFELY Collaborative, John Tazare, Alex J Walker, Laurie Tomlinson, George Hickman, Christopher T Rentsch, Elizabeth J Williamson, Krishnan Bhaskaran, David Evans, Kevin Wing, Rohini Mathur, Angel YS Wong, Anna Schultze, Seb Bacon, Chris Bates, Caroline E Morton, Helen J Curtis, Emily Nightingale, Helen I McDonald, Amir Mehrkar, Peter Inglesby, Simon Davy, Brian MacKenna, Jonathan Cockburn, William J Hulme, Charlotte Warren-Gash, Ketaki Bhate, Dorothea Nitsch, Emma Powell, Amy Mulick, Harriet Forbes, Caroline Minassian, Richard Croker, John Parry, Frank Hester, Sam Harper, Rosalind M Eggo, Stephen JW Evans, Liam Smeeth, Ian J Douglas, Ben Goldacre

## Abstract

**Background:** Patients with COVID-19 are thought to be at higher risk of cardiometabolic and pulmonary complications, but quantification of that risk is limited. We aimed to describe the overall burden of these complications in survivors of severe COVID-19.

**Methods:** Working on behalf of NHS England, we used linked primary care records, death certificate and hospital data from the OpenSAFELY platform. We constructed three cohorts: patients discharged following hospitalisation with COVID-19, patients discharged following hospitalisation with pneumonia in 2019, and a frequency-matched cohort from the general population in 2019. We studied eight cardiometabolic and pulmonary outcomes. Absolute rates were measured in each cohort and Cox regression models were fitted to estimate age/sex adjusted hazard ratios comparing outcome rates between discharged COVID-19 patients and the two comparator cohorts.

**Results:** Amongst the population of 31,716 patients discharged following hospitalisation with COVID-19, rates for majority of outcomes peaked in the first month post-discharge, then declined over the following four months. Patients in the COVID-19 population had markedly increased risk of all outcomes compared to matched controls from the 2019 general population, especially for pulmonary embolism (HR 12.86; 95% CI: 11.23 - 14.74). Outcome rates were more similar when comparing patients discharged with COVID-19 to those discharged with pneumonia in 2019, although COVID-19 patients had increased risk of type 2 diabetes (HR 1.23; 95% CI: 1.05 - 1.44).

**Interpretation:** Cardiometabolic and pulmonary adverse outcomes are markedly raised following hospitalisation for COVID-19 compared to the general population. However, the excess risks were more comparable to those seen following hospitalisation with pneumonia. Identifying patients at particularly high risk of outcomes would inform targeted preventive measures.

**Funding:** Wellcome, Royal Society, National Institute for Health Research, National Institute for Health Research Oxford Biomedical Research Centre, UK Medical Research Council, UK Research and Innovation, Health and Safety Executive.

## Introduction

Cardiometabolic and pulmonary complications, especially thrombotic events, have been described as a key feature of the severe acute phase of COVID-19. A recent systematic review estimated the risk of venous thromboembolism (VTE) to be ∼15% in hospitalised COVID-19 patients, with higher risks observed in people admitted to intensive care (∼30%^1–3^). Underlying reasons for this increased risk are likely to be multifactorial, including immobility following illness/hospitalisation as well as the known association with infection in general, mediated through interactions with general inflammatory and other immune pathways.^4^ The possibility that SARS-CoV-2 may directly trigger pulmonary thrombi via vascular damage and inflammatory effects in the lung has also been raised.^5^

As the COVID-19 pandemic has progressed, it is increasingly reported that some patients who recover from the acute disease phase go on to experience a range of post-recovery clinical problems. This post-acute COVID-19 syndrome is currently not well described or understood, with the UK National Institute for Health and Care Excellence stating that any body system could be affected, for an undetermined period of time.^6^ Any such syndrome now needs to be defined and quantified so that patients and health services can know what outcomes may be expected, and plan accordingly.^6,7^ It is also unclear whether COVID-19 is exceptional in its association with cardiometabolic events, or comparable to other respiratory pathogens, such as influenza and Streptococcus pneumoniae, which have well-described associations with acute cardiovascular events.

Work to date on cardiometabolic outcomes with COVID-19 has largely focused on risks during hospitalisation, with a lack of evidence on how these risks evolve in survivors of severe COVID-19. We therefore measured the rates of cardiometabolic outcomes in people in England with COVID-19, focusing on those who were discharged from hospital following the acute phase of COVID-19. For context, we compared these rates with those seen prior to the pandemic in both the general population and amongst people discharged following hospitalization for non-COVID-19 pneumonia.

## Methods

### Study design and data sources

We conducted an observational cohort study using electronic health record (EHR) data from primary care practices using TPP software linked to Office for National Statistics (ONS) death registrations and Secondary Uses Service (SUS) data (containing hospital records) through OpenSAFELY. This is a data analysis platform developed during the COVID-19 pandemic, on behalf of NHS England, to allow near real-time analysis of pseudonymised primary care records at scale, currently covering approximately 40% of the population in England, operating within the EHR vendor’s highly secure data centre.^8,9^ Details on Information Governance for the OpenSAFELY platform can be found in the Appendix.

### Population

We included all adults aged ≥18 years registered with a general practice for ≥1 year on the index date with information on age, sex, and socioeconomic deprivation. From this source population we selected three cohorts: all patients hospitalised with COVID-19 in 2020, a comparison cohort containing all patients hospitalised with non-COVID pneumonia across the equivalent period in 2019 and a general population frequency matched cohort in 2019. The COVID-19 and pneumonia cohorts were selected as anyone hospitalised with an associated diagnostic code for COVID-19 or pneumonia respectively (referred to as the “index hospitalisation”). The general population cohort was formed by matching each patient in the COVID-19 cohort to up to five patients eligible on 1st February 2019 in TPP on age (within 1 year), sex and region defined by Sustainability Transformation Partnership level (a more granular form of England NHS region).

### Outcomes and follow-up

We measured eight outcomes: deep vein thrombosis (DVT), pulmonary embolism (PE), ischaemic stroke, myocardial infarction (MI), heart failure, acute kidney injury (AKI) and new type 2 diabetes mellitus (T2DM) diagnosis.

The study periods ran between 1st February and 1st November in either 2019 or 2020, depending on the population (as defined above). The follow-up period began on the discharge date of the index COVID-19 or pneumonia hospital stay or 1st February 2019 in the general population matched cohort. For each analysis, follow-up ended on the earliest of: the first recorded outcome event, the study end date, or the date of death of the patient. For the AKI outcome, we excluded patients who were receiving dialysis before the index date (defined as presence of a dialysis code or eGFR < 15ml/min). For diabetes, we excluded any patients who had a previous diabetes event, to ensure only incident diagnoses were measured.

Outcomes were defined primarily as the presence of a diagnostic code for each of the respective outcomes, either in the general practice record, in hospital, or as a cause of death on a death certificate. For the primary analysis, we excluded use of a GP record if the patient had a recent outcome recorded within three months before the index date (including if they had been recorded during the index hospitalisation). This was to prevent double counting of the same event, for example where a GP updates the record of a patient, recording an event that occurred during a recent hospitalisation. Sensitivity analyses explored the effect of including these events (see Statistical Methods).

### Statistical Methods

We described the demographics of the three patient cohorts formed: patients discharged from an admission with COVID-19, patients discharged from an admission with pneumonia and patients from the general population matched on age, sex and region.

Rates were reported for each outcome, per 10,000 person months, initially for the whole follow-up time, then stratified into time windows: 0-29 days, 30-59 days, 60-89 days, 90-120 days and 120+ days post discharge for the COVID-19 and pneumonia cohorts, to determine how the rate of outcomes changed over time. Rates in all three patient cohorts were stratified by age, sex and ethnicity.

We used Cox regression models to estimate hazard ratios (HRs) and 95% confidence intervals (CIs) to compare the rates of each outcome between, 1) the discharged COVID-19 group and the matched general population group, and 2) the discharged COVID-19 group and the discharged pneumonia group. We investigated crude univariable and age and sex adjusted models. The same patient can contribute person-time to all exposure groups, however these periods are non-overlapping therefore we applied robust standard errors.

In sensitivity analyses we tested the effect of including 1) previously omitted outcomes recorded in the primary care record when there was a recorded outcome within 3 months before the index date, and 2) only events recorded in hospital or as a cause of death on a death certificate.

### Software and reproducibility

Data management was performed using the OpenSAFELY software, Python 3.8 and SQL, and analysis using Stata 16.1. All codelists alongside code for data management and analyses can be found at: https://github.com/opensafely/post-covid-outcomes-research. All software for the OpenSAFELY platform is available for review and re-use at https://github.com/opensafely.

## Results

We identified 31,716 patients discharged following an admission with COVID-19 in 2020 and 89,185 patients discharged with pneumonia in 2019. For each patient discharged following an admission with COVID-19 we matched up to five patients from the general population eligible in TPP on 1st February 2019 on age, sex and region. We successfully matched five patients for over 99.9% of COVID-19 patients.

Demographics for the cohorts studied are summarised in Table 1. Compared to the COVID-19 cohort, the pneumonia cohort had a higher proportion aged over 80. The ethnic breakdown was broadly similar between the three groups, although COVID-19 patients had a higher proportion of patients who were Asian and Asian British.

**Table 1:** Patient demographics amongst patients discharged with COVID-19 in 2020, all patients discharged with non-COVID pneumonia in 2019 and a matched (age, sex and region) general population comparator group in 2019

| Demographic (%) | Discharged<br>Pneumonia (2019) | Matched General<br>Population (2019) | Discharged<br>COVID-19 |
| --- | --- | --- | --- |
| Total | 89,185 | 158,551 | 31,716 |
| Age |  |  |  |
| 18 - 49 | 8,903 (10.0) | 32,390 (20.4) | 6,478 (20.4) |
| 50 - 59 | 8,136 (9.1) | 24,730 (15.6) | 4,946 (15.6) |
| 60 - 69 | 13,494 (15.1) | 26,165 (16.5) | 5,233 (16.5) |
| 70 - 79 | 22,912 (25.7) | 31,769 (20.0) | 6,354 (20.0) |
| 80 + | 35,740 (40.1) | 43,497 (27.4) | 8,705 (27.4) |
| Sex |  |  |  |
| Male | 44,257 (49.6) | 75,062 (47.3) | 15,016 (47.3) |
| Female | 44,928 (50.4) | 83,489 (52.7) | 16,700 (52.7) |
| Ethnicity |  |  |  |
| White | 59,636 (66.9) | 102,274 (64.5) | 19,422 (61.2) |
| Mixed | 327 (0.4) | 1,058 (0.7) | 307 (1.0) |
| Asian /Asian British | 3,202 (3.6) | 8,376 (5.3) | 3,323 (10.5) |
| Black | 844 (0.9) | 2,761 (1.7) | 1,048 (3.3) |
| Other | 506 (0.6) | 1,967 (1.2) | 516 (1.6) |
| Unknown | 24,670 (27.7) | 42,115 (26.6) | 7,100 (22.4) |

Overall rates of each outcome per 10,000 person months for the whole follow-up are presented in Table 2. For the majority of outcomes we observed higher rates of serious cardiometabolic and pulmonary complications in discharged pneumonia patients compared to both discharged COVID-19 patients and the matched general population group (Table 2, Figure 1). Across all three cohorts, the largest absolute rates were for AKI and heart failure. Overall rates stratified by age, sex and ethnicity are presented in the appendix (Tables A5-A12). For COVID-19 patients, rates of stroke and T2DM were consistently higher amongst the over 80 compared to the other comparison groups, although the pattern was not consistent across other ages groups. Similarly, rates were not constant by ethnic group, for example, the rate of new T2DM diagnoses were slightly higher amongst black patients discharged following an admission with COVID-19.

**Table 2:** Overall rates of outcomes (events per 10,000 person months) amongst patients discharged with COVID-19 in 2020, all patients discharged with non-COVID pneumonia in 2019 and a matched (age, sex and region) general population comparator group in 2019.

| Outcome | Analysis Group | Person time<br>(x 10,000<br>person months) | Number of<br>events | Rate (95% CI) |
| --- | --- | --- | --- | --- |
| Stroke | Discharged COVID-19 | 13.6 | 544 | 40.1 (36.9-43.7) |
|  | General Population | 139.5 | 1140 | 8.2 (7.7-8.7) |
|  | Discharged Pneumonia | 36.4 | 1888 | 51.8 (49.6-54.2) |
| DVT | Discharged COVID-19 | 13.6 | 261 | 19.3 (17.1-21.7) |
|  | General Population | 139.6 | 430 | 3.1 (2.8-3.4) |
|  | Discharged Pneumonia | 36.5 | 804 | 22.0 (20.6-23.6) |
| PE | Discharged COVID-19 | 13.5 | 521 | 38.6 (35.4-42.1) |
|  | General Population | 139.7 | 379 | 2.7 (2.5-3.0) |
|  | Discharged Pneumonia | 36.3 | 1528 | 42.1 (40.0-44.2) |
| Heart Failure | Discharged COVID-19 | 13 | 2621 | 202.3 (194.7-210.2) |
|  | General Population | 138 | 4625 | 33.5 (32.6-34.5) |
|  | Discharged Pneumonia | 33.5 | 12181 | 363.8 (357.4-370.4) |
| MI | Discharged COVID-19 | 13.6 | 297 | 21.9 (19.5-24.5) |
|  | General Population | 139.5 | 858 | 6.1 (5.8-6.6) |
|  | Discharged Pneumonia | 36.5 | 1251 | 34.3 (32.5-36.3) |
| AKI | Discharged COVID-19 | 12.9 | 2081 | 161.4 (154.6-168.5) |
|  | General Population | 138.5 | 3154 | 22.8 (22.0-23.6) |
|  | Discharged Pneumonia | 34.3 | 7817 | 228.1 (223.1-233.2) |
| T2DM | Discharged COVID-19 | 9.2 | 293 | 31.7 (28.3-35.6) |
|  | General Population | 115.2 | 886 | 7.7 (7.2-8.2) |
|  | Discharged Pneumonia | 25.8 | 848 | 32.9 (30.8-35.2) |

**Figure 1:**
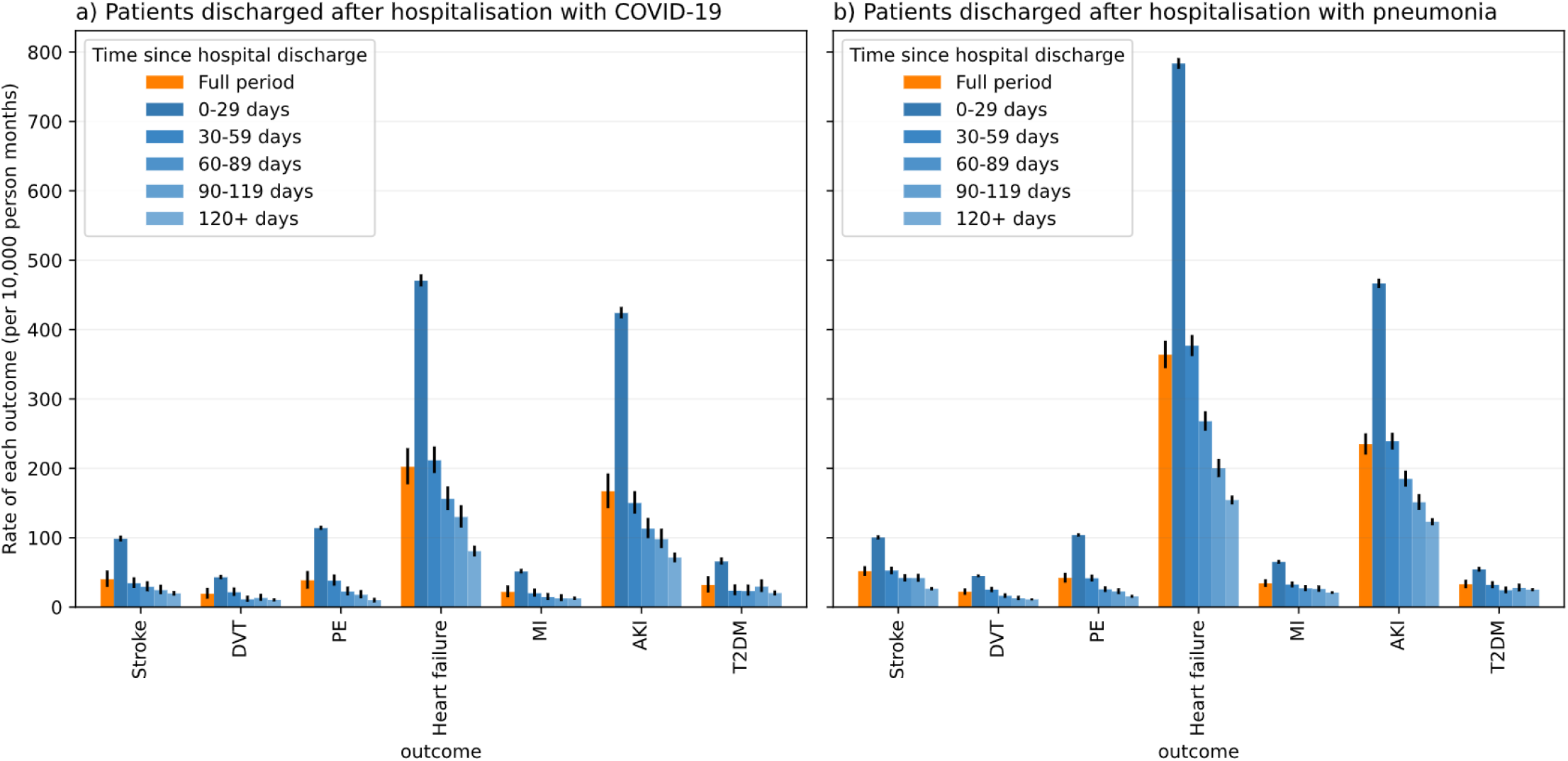
Rate of outcomes (events per 10,000 person months) in time periods following hospital discharge for COVID-19/pneumonia.

Stratified overall rates in 30-day time windows are shown in Figure 1. For discharged COVID-19 patients, we observed the highest rates for all outcomes in the first 30 days post-discharge, with a gradual decline in subsequent periods, consistent with the pattern of rates observed for patients discharged with pneumonia in 2019. For the discharged COVID-19 and pneumonia groups, we observed pronounced rates of AKI and heart failure in the first 30 days post-discharge (Figure 1).

After age and sex adjustment, the discharged COVID-19 group had markedly increased risk for all outcomes compared to the matched general population group (Table 3, Figure 2). The largest increase in risk was observed for PE (HR 12.86; 95% CI: 11.23 - 14.74) and AKI (HR 6.72; 95% CI: 6.35 - 7.12). For the comparison with discharged pneumonia patients, discharged COVID-19 patients had similar risks for the majority of outcomes. We observed an increased risk of new T2DM in patients discharged with COVID-19 compared to patients discharged with pneumonia (HR 1.23; 95% CI: 1.05 - 1.44).

**Table 3:** Estimated hazard ratios from univariable and age and sex adjusted Cox regression models comparing the risk of outcomes in patients who were hospitalised with COVID-19 and then discharged, compared to 1) patients who were hospitalised with pneumonia and then discharged in 2019, and 2) an age, sex and region matched general population comparator group in 2019. The number of events are measured per 10,000 person months.

| Outcome | Comparator | Comparator |  | Discharged COVID-19 |  | Crude HR (95% CI) | Age/Sex adjusted HR (95% CI) |
| --- | --- | --- | --- | --- | --- | --- | --- |
|  |  | Person Time | Number of events | Person Time | Number of events |  |  |
| Stroke | General Population | 139.5 | 1140 | 13.6 | 544 | 4.64 (4.18-5.15) | 4.92 (4.43-5.47) |
|  | Pneumonia | 36.4 | 1888 | 13.6 | 544 | 0.79 (0.72-0.87) | 0.96 (0.84-1.09) |
| DVT | General Population | 139.6 | 430 | 13.6 | 261 | 5.91 (5.04-6.92) | 6.09 (5.20-7.13) |
|  | Pneumonia | 36.5 | 804 | 13.6 | 261 | 0.89 (0.78-1.03) | 0.87 (0.73-1.03) |
| PE | General Population | 139.7 | 379 | 13.5 | 521 | 12.61 (11.00-14.45) | 12.86 (11.23-14.74) |
|  | Pneumonia | 36.3 | 1528 | 13.5 | 521 | 0.96 (0.87-1.06) | 0.96 (0.85-1.08) |
| Heart Failure | General Population | 138 | 4625 | 13 | 2621 | 5.08 (4.83-5.33) | 5.52 (5.25-5.80) |
|  | Pneumonia | 33.5 | 12181 | 13 | 2621 | 0.58 (0.56-0.61) | 0.80 (0.75-0.84) |
| MI | General Population | 139.5 | 858 | 13.6 | 297 | 3.31 (2.89-3.78) | 3.47 (3.04-3.97) |
|  | Pneumonia | 36.5 | 1251 | 13.6 | 297 | 0.65 (0.58-0.74) | 0.89 (0.75-1.05) |
| AKI | General Population | 138.5 | 3154 | 12.9 | 2081 | 6.29 (5.94-6.66) | 6.72 (6.35-7.12) |
|  | Pneumonia | 34.3 | 7817 | 12.9 | 2081 | 0.73 (0.70-0.77) | 0.90 (0.85-0.96) |
| T2DM | General Population | 115.2 | 886 | 9.2 | 293 | 3.86 (3.37-4.42) | 3.96 (3.46-4.53) |
|  | Pneumonia | 25.8 | 848 | 9.2 | 293 | 1.00 (0.87-1.14) | 1.23 (1.05-1.44) |

**Figure 2:**
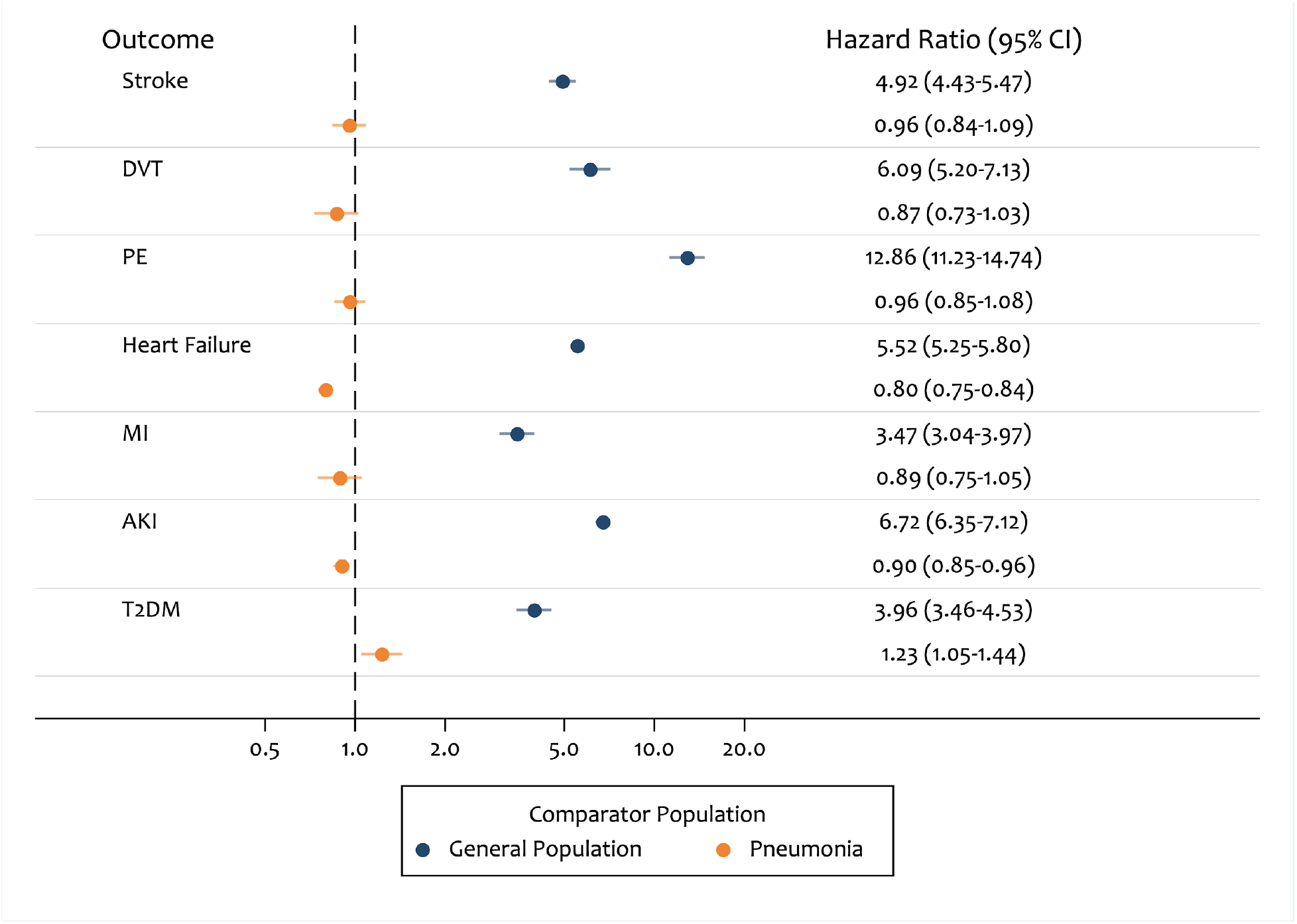
Age and sex adjusted hazard ratios for risk of outcomes in patients who were hospitalised with COVID-19 and then discharged, compared to 1) patients who were hospitalised with pneumonia and then discharged in 2019, and 2) an age, sex and region matched general population comparator group in 2019.

Results from a sensitivity analysis investigating robustness of absolute and relative rates to outcome definition are presented in the Appendix. Change in outcome definitions did not meaningfully alter conclusions.

## Discussion

### Key findings

In this descriptive study, we set out to report the overall rates of eight cardiometabolic and pulmonary outcomes in three cohorts: 1) patients discharged following hospitalisation with COVID-19, 2) patients discharged following hospitalisation with pneumonia in 2019 and 3) a frequency-matched group of patients from the general population in 2019. We found that the rate of cardiometabolic and pulmonary complications following discharge from hospitalisation with COVID-19 was notably higher compared to an age, sex and region matched general population cohort, especially for PE and AKI. However patients discharged with COVID-19 followed a broadly similar pattern of elevated rates to those discharged from hospital after pneumonia in 2019. The pattern of change in the rate of outcomes over time following discharge from hospital was also broadly similar between the COVID-19 and pneumonia patients, with the highest rate in the initial 30 days of follow-up, then a 2-3 fold drop in the next 30 days, followed by a more gradual decline. In both discharge based cohorts, rates remained substantial even after more than 120 days.

### Strengths and limitations

We were able to source our cohorts from the OpenSAFELY platform, which contains data on over 17m adults. This gave us a population who were discharged following hospitalisation with COVID-19 of 31,716, allowing us to obtain precise estimates of the rate of each outcome. We were also able to draw on multiple linked data sources, including primary care records, hospitalisations and death certificates. This allows a more complete picture to be presented of the clinical activity surrounding each outcome.

We believe that our use of both a general population and active control population of patients hospitalised with pneumonia in 2019 provides useful context for the rates of these outcomes in COVID-19 patients who survive hospitalisation. Presenting the rates in this context is more informative than within a general population alone and offers an important comparison with a cohort experiencing exposure to another acute respiratory illness event requiring hospitalisation.

Our study aimed to describe clinical events that occurred *after* discharge from hospital, and not the total additional morbidity burden of COVID-19 hospitalisation: specifically we did not set out to describe events that occurred *during* hospital admission with COVID-19 or pneumonia. However, in our view reliable analysis of in-hospital events may only be achievable with bespoke collections of detailed hospital data, due to shortcomings in routinely collected administrative data that are widely used for such analyses. For example, SUS and HES data contain a list of diagnostic codes associated with each hospitalisation, but they do not contain sufficient information to determine the exact timing of all events within each hospitalisation episode. This means that time-to event analyses are not possible. Similarly it is not possible to reliably determine the sequence of events during hospitalisation: so a patient hospitalised with COVID-19, who later had a stroke, may be coded in a similar way to a patient who was hospitalised with a stroke, and then infected with SARS-CoV-2 while in hospital. In addition, routine PCR testing on hospitalised patients during the pandemic may lead to very high ascertainment of infection with SARS-CoV-2, which may not have occurred to the same extent in the comparison population for pneumonia admissions.

We did not attempt to determine here whether any observed differences were due to a particular feature of the pathophysiology associated with SARS-CoV-2 infection, or whether other factors might have had a greater influence, such as competing risk of death, pre-existing patient comorbidities, changes in healthcare provision during the pandemic or differing likelihood of ascertainment of pre-existing conditions. In the context of the rapidly changing pandemic, we aimed to provide an overview of the rates of outcomes after discharge from hospitalisation with COVID-19, compared with pneumonia, to inform health services.

It has been reported that there was a marked reduction in GP and hospital activity during the first wave of the pandemic, for example a 40% reduction in admissions for acute coronary syndrome.^10–12^ This may be in part explained by a reluctance of patients to present at healthcare services for fear of contracting the virus. As a result, we believe population-level rates of many outcomes will be under-ascertained during 2020 compared with 2019. It is unknown whether this applies in the same way to patients who have already had severe COVID-19; if ascertainment is lower, then this would result in a possible under-estimate of outcome rates associated with COVID-19 in our study.

### Research in context

A recent observational study measured similar outcome events in a population of patients discharged from hospital following COVID-19.^13^ They observed elevated rates in the COVID-19 population compared to a matched general population control group. Our findings are consistent in showing similarly increased rates of outcomes in patients post-discharge with COVID-19. However, importantly we further show that these increased rates of outcomes are broadly comparable, if not slightly lower, when compared to people discharged from hospital following pneumonia, selected as a major non-COVID respiratory infection.

The impact of the post COVID-19 hospitalisation events described in this study upon the NHS in England is substantial. Future work should also investigate any association between non-hospitalised SARS-CoV-2 infection and these outcomes, and quantify any likely population level impact.

### Summary

The rate of cardiometabolic and pulmonary events in COVID-19 survivors discharged from hospitalisation was elevated in a similar manner to patients discharged from hospitalisation with pre-pandemic pneumonia. Next steps include seeing whether patients at highest risk of post-covid outcomes can be identified and determining whether higher risk groups could be early targeted for possible preventative action.

## Data Availability

All data were linked, stored and analysed securely within the OpenSAFELY platform (https://opensafely.org/). Detailed pseudonymized patient data are potentially re-identifiable and therefore not shared. We rapidly delivered the OpenSAFELY data analysis platform without prior funding to deliver timely analyses of urgent research questions in the context of the global COVID-19 health emergency: now that the platform is established, we are developing a formal process for external users to request access in collaboration with NHS England. Details of this process will be published in due course on the OpenSAFELY website.

## Acknowledgements

We are very grateful for all the support received from the TPP Technical Operations team throughout this work; for generous assistance from the information governance and database teams at NHS England / NHSX.

## Ethical Approval

This study was approved by the Health Research Authority (REC 20/LO/0651) and by the LSHTM Ethics Board (#21863).

## Contributorship

B.G. conceived the OpenSAFELY platform and the approach. BG and LS led the project overall and are guarantors. JT, AJW, and IJD designed the study, conducted the statistical analysis and wrote the first draft. SB and GH led on software development. AM led on information governance. SB, CEM, DE, SD, GH, PI, JC, AJW, GH, JC, DE, WJH and FH. contributed to software development. JT, AW, LT, ID, KW, RM conceptualised disease categories and code lists. JT and AJW wrote statistical analysis code. EJW, HJC, LS, and BG obtained ethical approvals. All authors were involved in design and conceptual development and reviewed and approved the final manuscript.

## Information governance and ethics

NHS England is the data controller; TPP is the data processor; and the key researchers on OpenSAFELY are acting on behalf of NHS England. OpenSAFELY is hosted within the TPP environment which is accredited to the ISO 27001 information security standard and is NHS IG Toolkit compliant;^14,15^ patient data are pseudonymised for analysis and linkage using industry standard cryptographic hashing techniques; all pseudonymised datasets transmitted for linkage onto OpenSAFELY are encrypted; access to the platform is via a virtual private network (VPN) connection, restricted to a small group of researchers who hold contracts with NHS England and only access the platform to initiate database queries and statistical models. Pseudonymised structured data include demographics, medications prescribed from primary care, diagnoses, and laboratory measures. No free text data are included. All database activity is logged; only aggregate statistical outputs leave the platform environment following best practice for anonymisation of results such as statistical disclosure control for low cell counts.^16^ The OpenSAFELY research platform adheres to the obligations of the UK General Data Protection Regulation (GDPR) and the Data Protection Act 2018. In March 2020, the Secretary of State for Health and Social Care used powers under the UK Health Service (Control of Patient Information) Regulations 2002 (COPI) to require organisations to process confidential patient information for the purposes of protecting public health, providing healthcare services to the public and monitoring and managing the COVID-19 outbreak and incidents of exposure; this sets aside the requirement for patient consent.^17^ Taken together, these provide the legal bases to link patient datasets on the OpenSAFELY platform. GP practices, from which the primary care data are obtained, are required to share relevant health information to support the public health response to the pandemic, and have been informed of the OpenSAFELY analytics platform. This study was approved by the Health Research Authority (REC reference 20/LO/0651) and by the LSHTM Ethics Board (ref 21863).

## Appendix Sensitivity analyses

**Figure A1:**
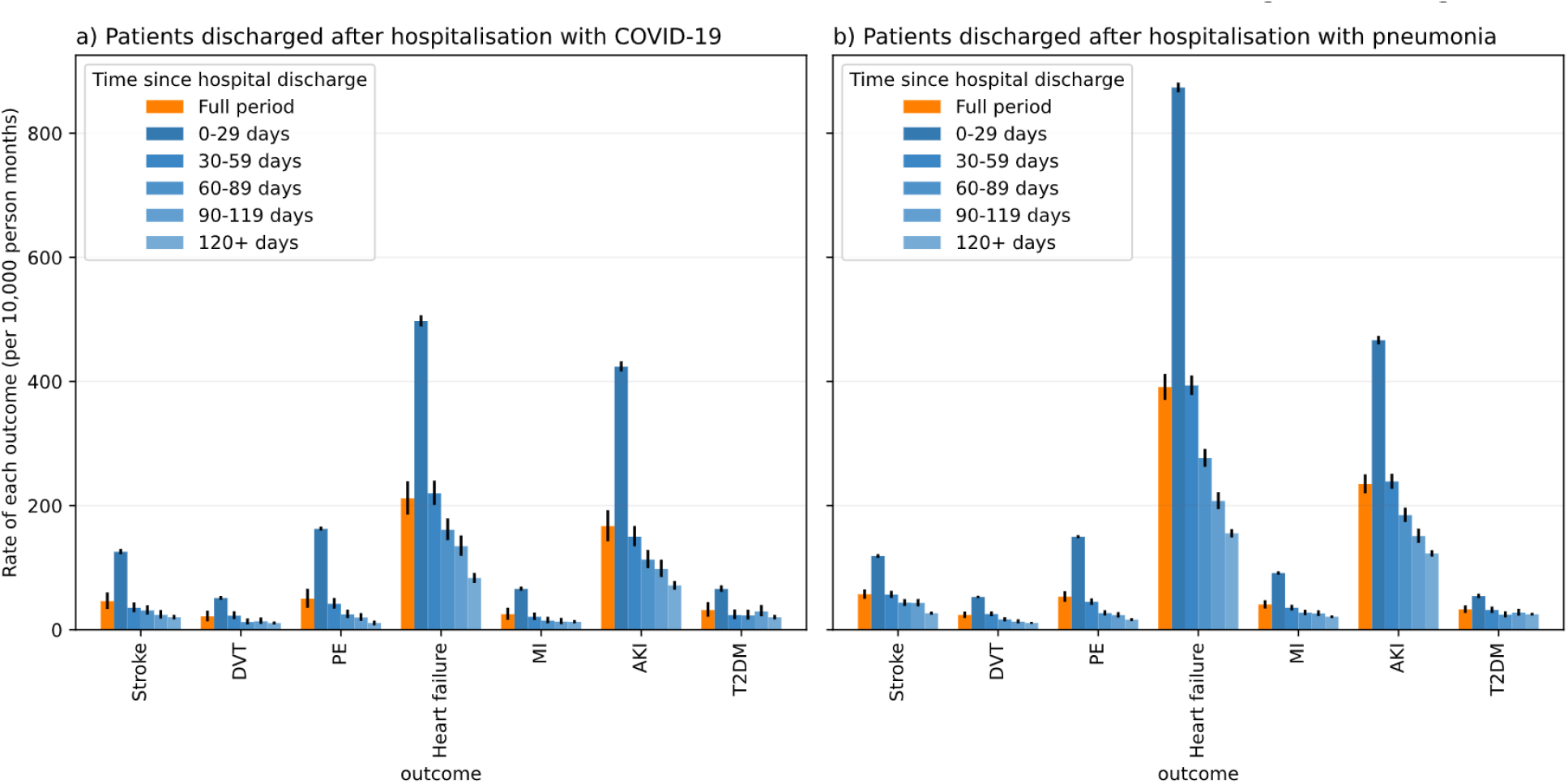
As Figure 1 but including all GP outcomes rather than censoring if the patient had an outcome within the 3 months before index date. AKI and diabetes are unchanged from figure 1.

**Figure A2:**
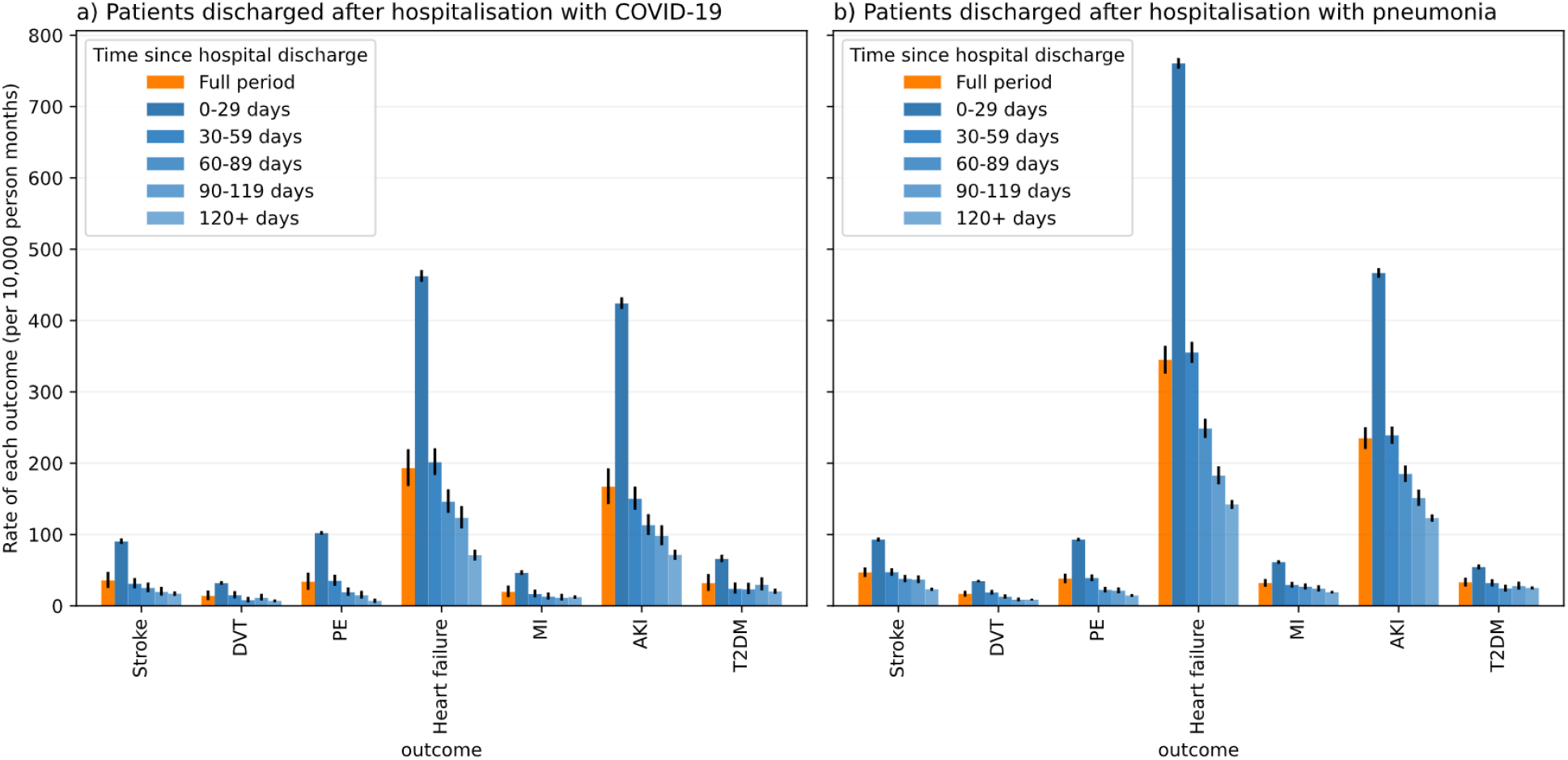
As Figure 1, but without using GP as an outcome (only hospitalisations and death certificate records). AKI and diabetes are unchanged from figure 1.

**Figure A3:**
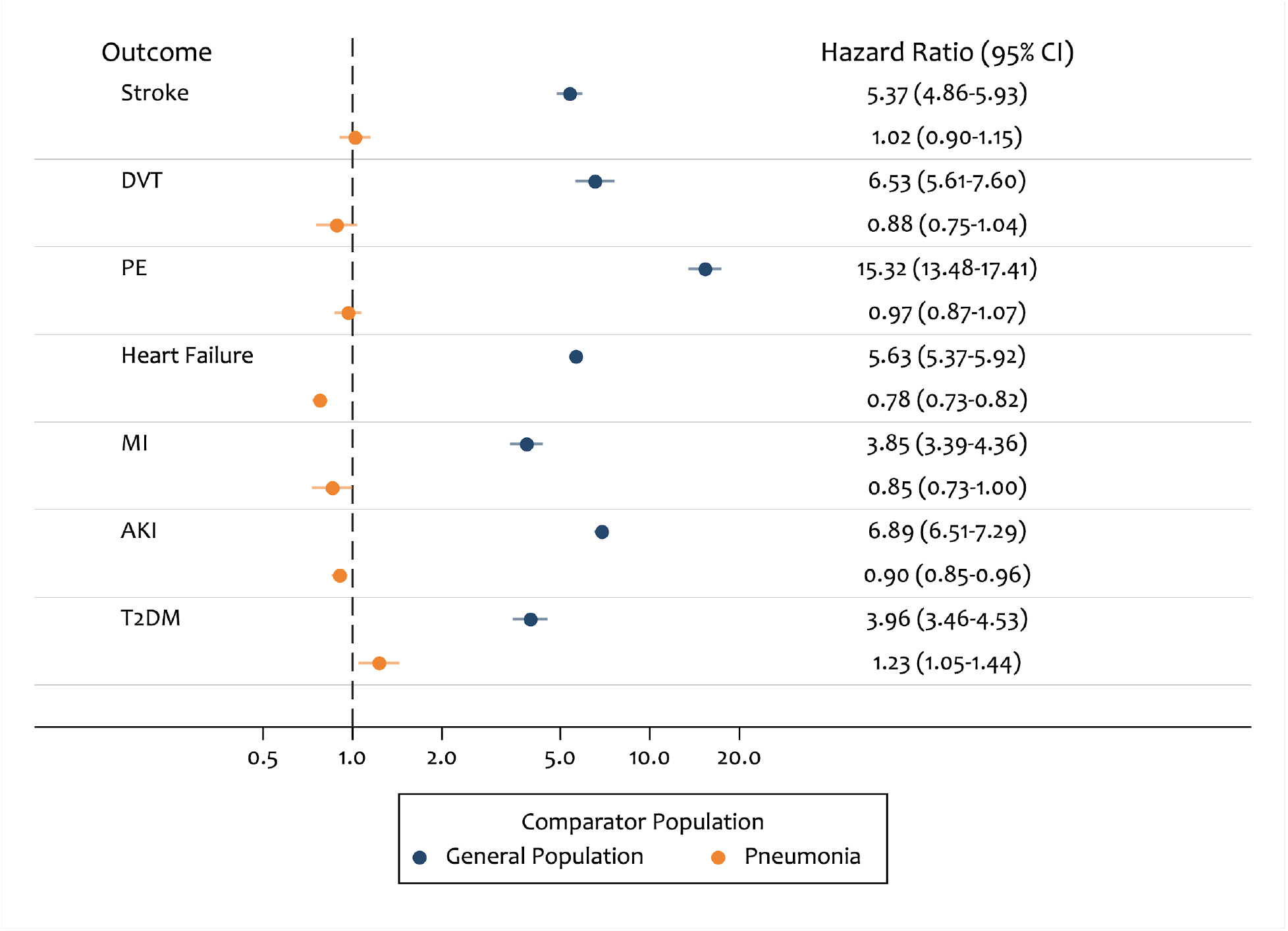
As Figure 2, but outcomes recorded in GP records are not censored if the patient had an outcome within the 3 months before index date.

**Figure A4:**
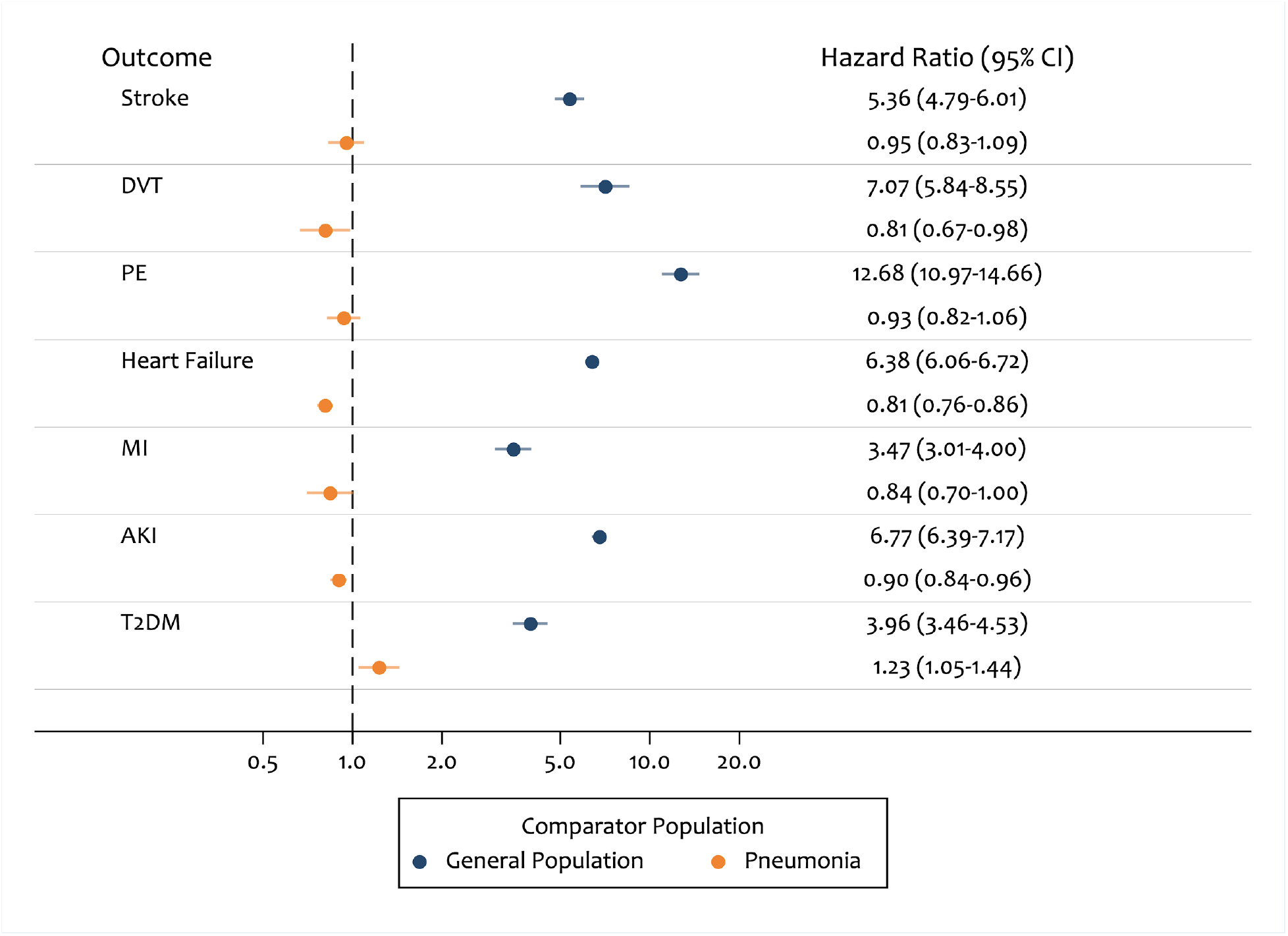
As Figure 2, but without using GP as an outcome (only hospitalisations and death certificate records).

**Table A1:** As Table 2, but outcomes recorded in GP records are not censored if the patient had an outcome within the 3 months before index date.

| Outcome | Analysis Group | Person Time (x<br>10,000 person<br>months) | Number of<br>events | Rate (95% CI) |
| --- | --- | --- | --- | --- |
| Stroke | Discharged COVID-19 | 13.5 | 624 | 46.2 (42.7-49.9) |
|  | General Population | 139.5 | 1167 | 8.4 (7.9-8.9) |
|  | Discharged Pneumonia | 36.3 | 2077 | 57.1 (54.7-59.7) |
| DVT | Discharged COVID-19 | 13.5 | 292 | 21.6 (19.2-24.2) |
|  | General Population | 139.6 | 440 | 3.2 (2.9-3.5) |
|  | Discharged Pneumonia | 36.5 | 872 | 23.9 (22.4-25.6) |
| PE | Discharged COVID-19 | 13.4 | 672 | 50.1 (46.4-54.0) |
|  | General Population | 139.7 | 392 | 2.8 (2.5-3.1) |
|  | Discharged Pneumonia | 36.2 | 1932 | 53.4 (51.1-55.8) |
| Heart Failure | Discharged COVID-19 | 12.9 | 2734 | 211.8 (204.0-219.9) |
|  | General Population | 137.9 | 4718 | 34.2 (33.2-35.2) |
|  | Discharged Pneumonia | 33.2 | 12966 | 391.1 (384.4-397.9) |
| MI | Discharged COVID-19 | 13.6 | 341 | 25.1 (22.6-28.0) |
|  | General Population | 139.5 | 872 | 6.3 (5.8-6.7) |
|  | Discharged Pneumonia | 36.4 | 1482 | 40.7 (38.7-42.9) |
| AKI | Discharged COVID-19 | 12.9 | 2148 | 167.0 (160.1-174.2) |
|  | General Population | 138.5 | 3162 | 22.8 (22.0-23.6) |
|  | Discharged Pneumonia | 34.2 | 8030 | 234.9 (229.8-240.1) |
| T2DM | Discharged COVID-19 | 9.2 | 293 | 31.7 (28.3-35.6) |
|  | General Population | 115.2 | 886 | 7.7 (7.2-8.2) |
|  | Discharged Pneumonia | 25.8 | 848 | 32.9 (30.8-35.2) |

**Table A2:** As Table 2, but without using GP as an outcome (only hospitalisations and death) certificate records.

| Outcome | Analysis Group | Person Time (x 10,000<br>person months) | Number of events | Rate (95% CI) |
| --- | --- | --- | --- | --- |
| Stroke | Discharged COVID-19 | 13.6 | 483 | 35.6 (32.5-38.9) |
|  | General Population | 139.6 | 925 | 6.6 (6.2-7.1) |
|  | Discharged Pneumonia | 36.5 | 1709 | 46.9 (44.7-49.1) |
| DVT | Discharged COVID-19 | 13.6 | 190 | 14.0 (12.1-16.1) |
|  | General Population | 139.7 | 266 | 1.9 (1.7-2.1) |
|  | Discharged Pneumonia | 36.5 | 613 | 16.8 (15.5-18.2) |
| PE | Discharged COVID-19 | 13.5 | 456 | 33.7 (30.8-37.0) |
|  | General Population | 139.7 | 336 | 2.4 (2.2-2.7) |
|  | Discharged Pneumonia | 36.4 | 1389 | 38.2 (36.2-40.2) |
| Heart Failure | Discharged COVID-19 | 13 | 2507 | 193.0 (185.6-200.7) |
|  | General Population | 138.4 | 3803 | 27.5 (26.6-28.4) |
|  | Discharged Pneumonia | 33.7 | 11610 | 344.8 (338.6-351.2) |
| MI | Discharged COVID-19 | 13.6 | 267 | 19.6 (17.4-22.1) |
|  | General Population | 139.6 | 775 | 5.6 (5.2-6.0) |
|  | Discharged Pneumonia | 36.5 | 1168 | 32.0 (30.2-33.9) |
| AKI | Discharged COVID-19 | 12.9 | 2051 | 158.9 (152.2-166.0) |
|  | General Population | 138.6 | 3076 | 22.2 (21.4-23.0) |
|  | Discharged Pneumonia | 34.3 | 7724 | 225.2 (220.2-230.3) |
| T2DM | Discharged COVID-19 | 9.2 | 293 | 31.7 (28.3-35.6) |
|  | General Population | 115.2 | 886 | 7.7 (7.2-8.2) |
|  | Discharged Pneumonia | 25.8 | 848 | 32.9 (30.8-35.2) |

**Table A3:**
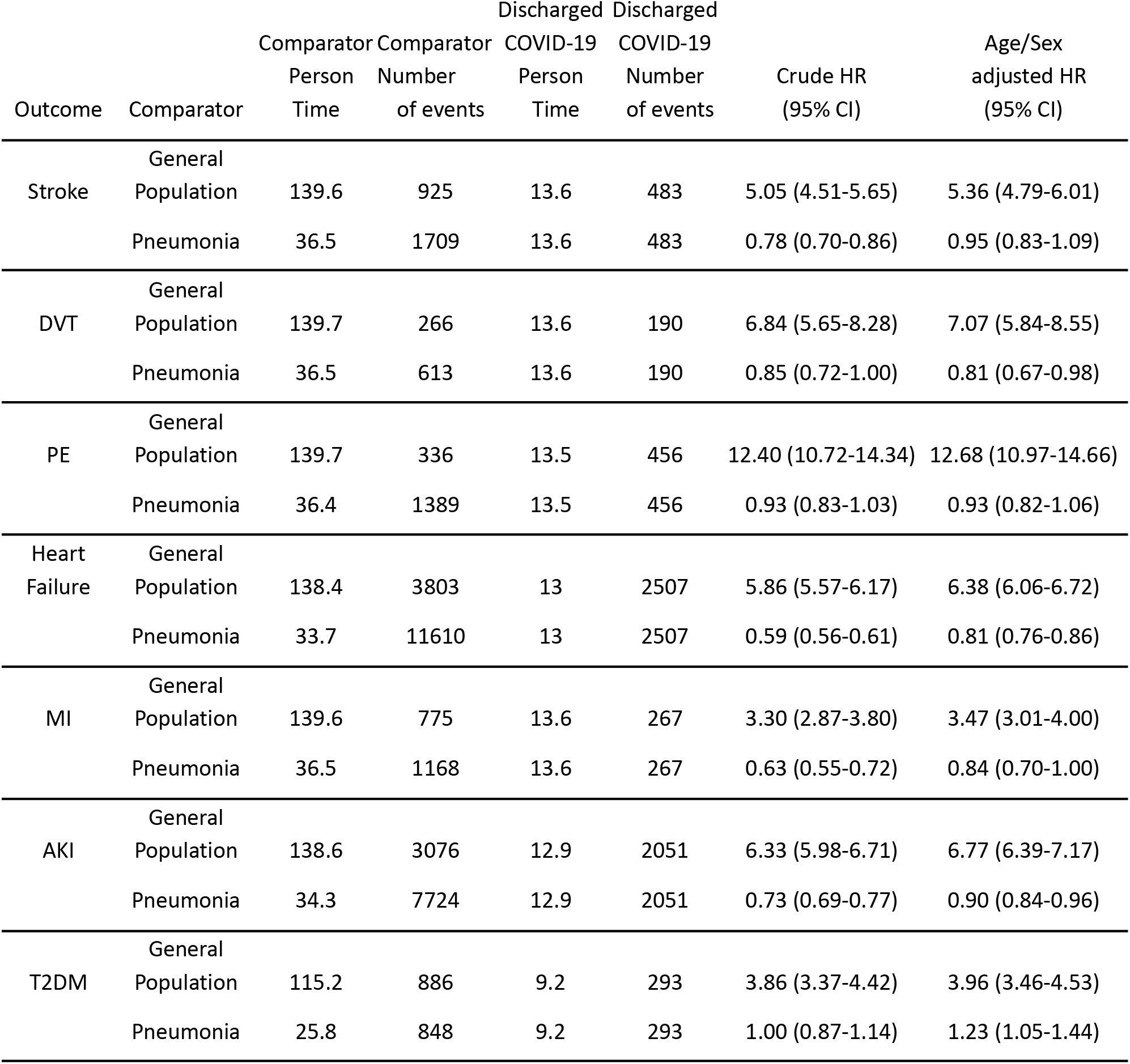
As Table 3, but without using GP as an outcome (only hospitalisations and death) certificate records.

| Outcome | Comparator | Comparator |  | Discharged COVID-19 |  | Crude HR<br>(95% CI) | Age/Sex<br>adjusted HR<br>(95% CI) |
| --- | --- | --- | --- | --- | --- | --- | --- |
|  |  | Person<br>Time | Number<br>of events | Person<br>Time | Number<br>of events |  |  |
| Stroke | General<br>Population | 139.6 | 925 | 13.6 | 483 | 5.05 (4.51-5.65) | 5.36 (4.79-6.01) |
|  | Pneumonia | 36.5 | 1709 | 13.6 | 483 | 0.78 (0.70-0.86) | 0.95 (0.83-1.09) |
| DVT | General<br>Population | 139.7 | 266 | 13.6 | 190 | 6.84 (5.65-8.28) | 7.07 (5.84-8.55) |
|  | Pneumonia | 36.5 | 613 | 13.6 | 190 | 0.85 (0.72-1.00) | 0.81 (0.67-0.98) |
| PE | General<br>Population | 139.7 | 336 | 13.5 | 456 | 12.40 (10.72-14.34) | 12.68 (10.97-14.66) |
|  | Pneumonia | 36.4 | 1389 | 13.5 | 456 | 0.93 (0.83-1.03) | 0.93 (0.82-1.06) |
| Heart<br>Failure | General<br>Population | 138.4 | 3803 | 13 | 2507 | 5.86 (5.57-6.17) | 6.38 (6.06-6.72) |
|  | Pneumonia | 33.7 | 11610 | 13 | 2507 | 0.59 (0.56-0.61) | 0.81 (0.76-0.86) |
| MI | General<br>Population | 139.6 | 775 | 13.6 | 267 | 3.30 (2.87-3.80) | 3.47 (3.01-4.00) |
|  | Pneumonia | 36.5 | 1168 | 13.6 | 267 | 0.63 (0.55-0.72) | 0.84 (0.70-1.00) |
| AKI | General<br>Population | 138.6 | 3076 | 12.9 | 2051 | 6.33 (5.98-6.71) | 6.77 (6.39-7.17) |
|  | Pneumonia | 34.3 | 7724 | 12.9 | 2051 | 0.73 (0.69-0.77) | 0.90 (0.84-0.96) |
| T2DM | General<br>Population | 115.2 | 886 | 9.2 | 293 | 3.86 (3.37-4.42) | 3.96 (3.46-4.53) |
|  | Pneumonia | 25.8 | 848 | 9.2 | 293 | 1.00 (0.87-1.14) | 1.23 (1.05-1.44) |

**Table A4:**
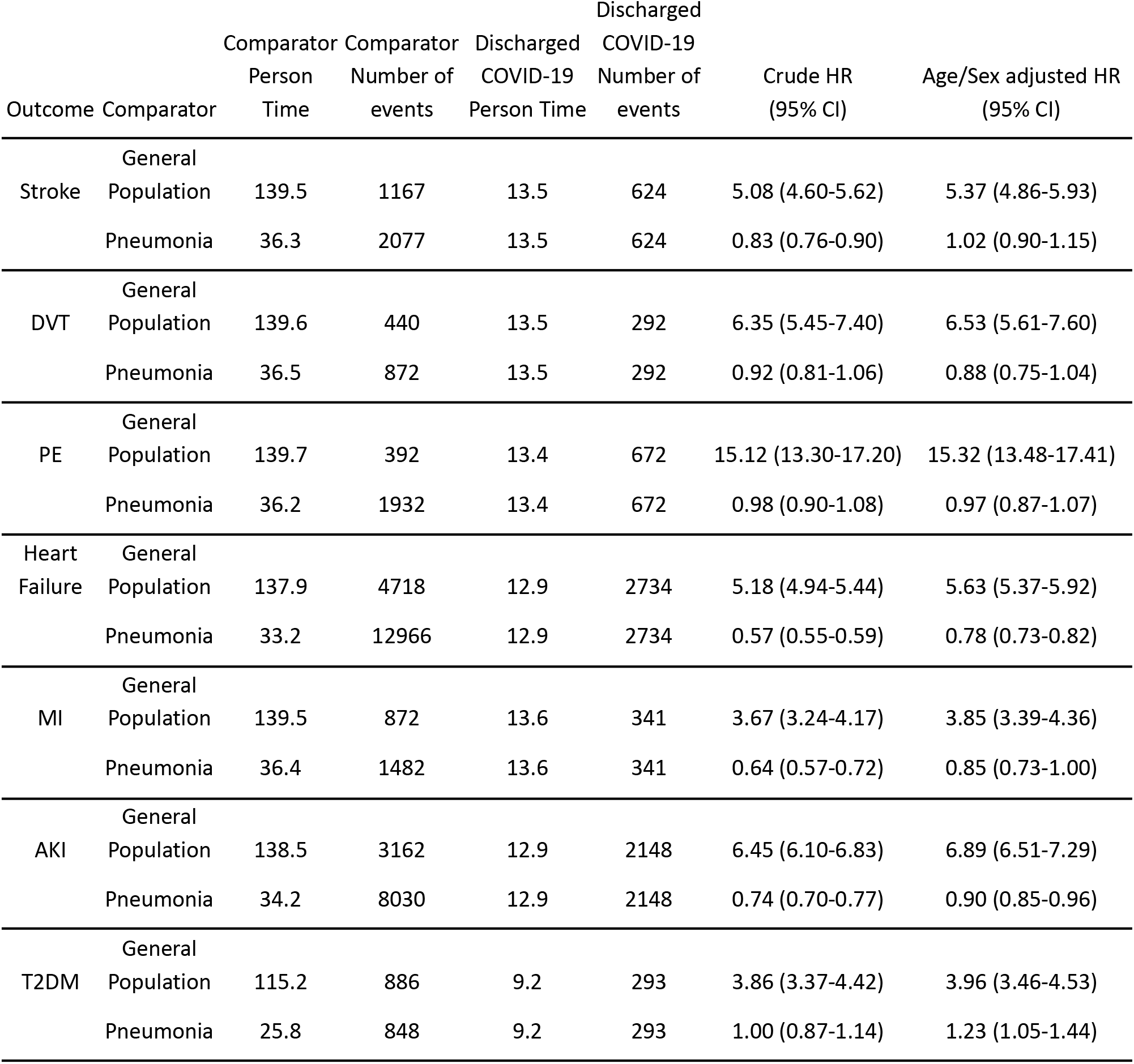
As Table 3, but outcomes recorded in GP records are not censored if the patient had an outcome within the 3 months before index date.

**Table A5:**
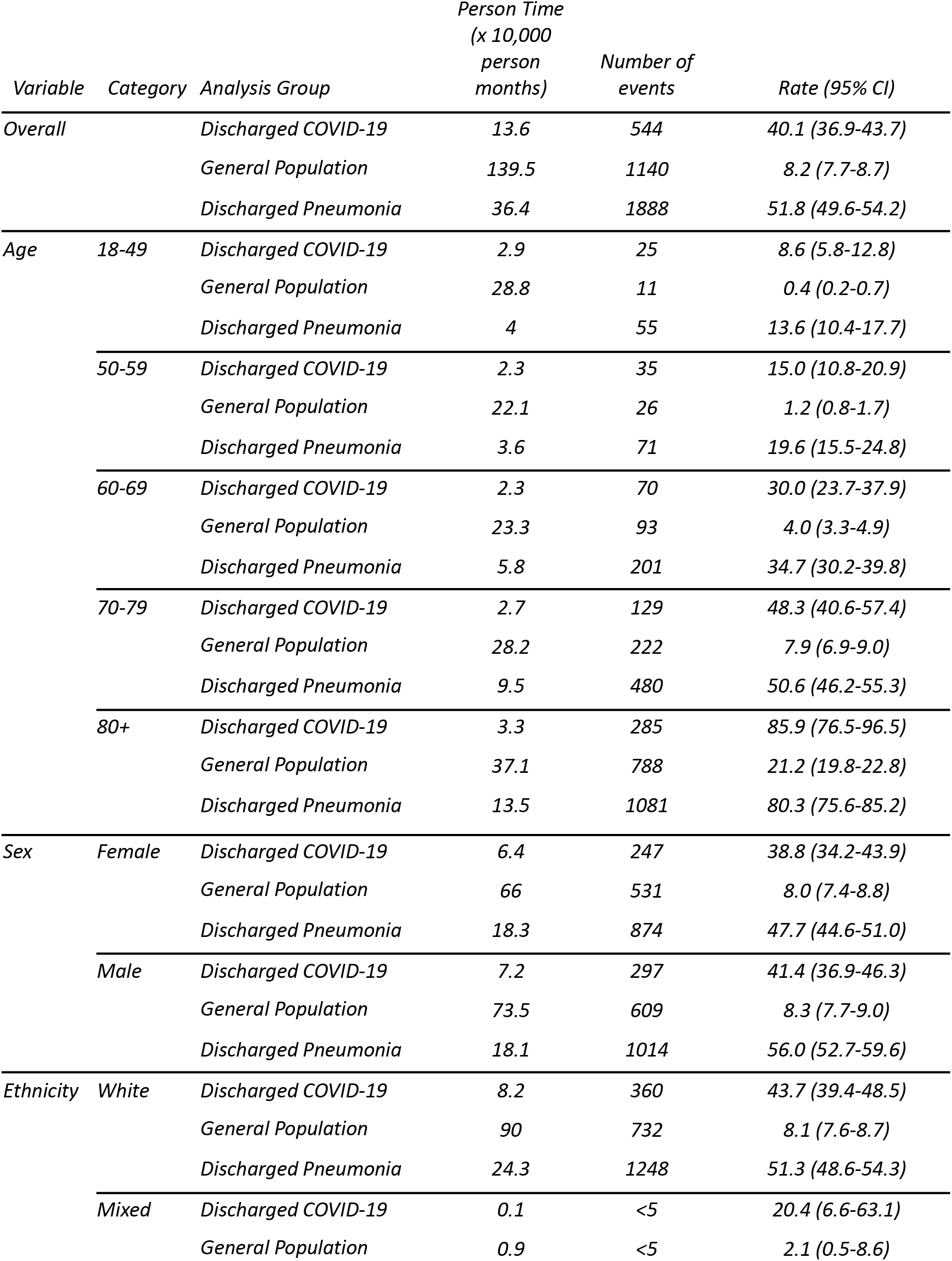
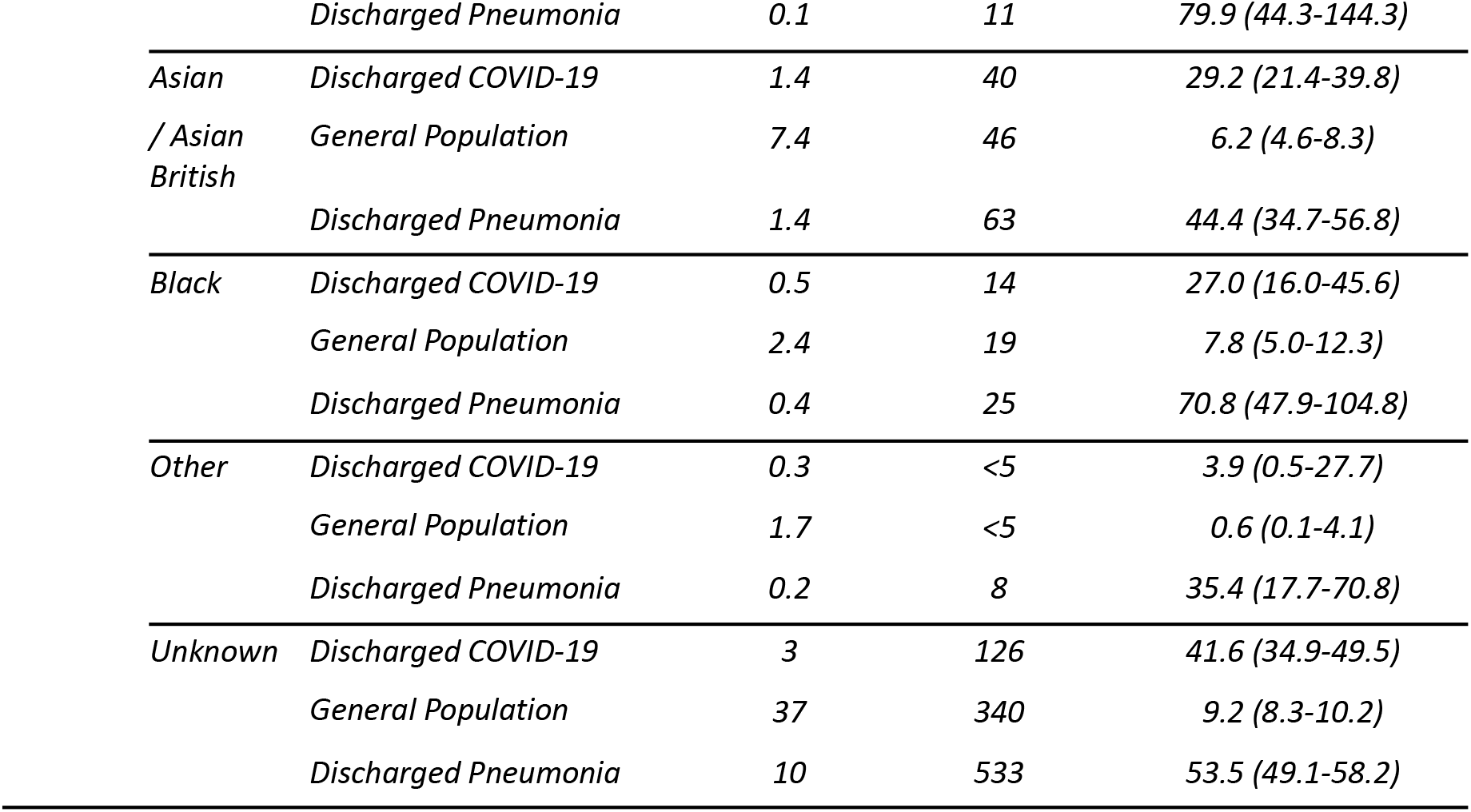
Rates of stroke amongst patients discharged with COVID-19 in 2020, all patients discharged with non-COVID pneumonia in 2019 and a matched (age, sex and region) general population comparator group in 2019

**Table A6:**
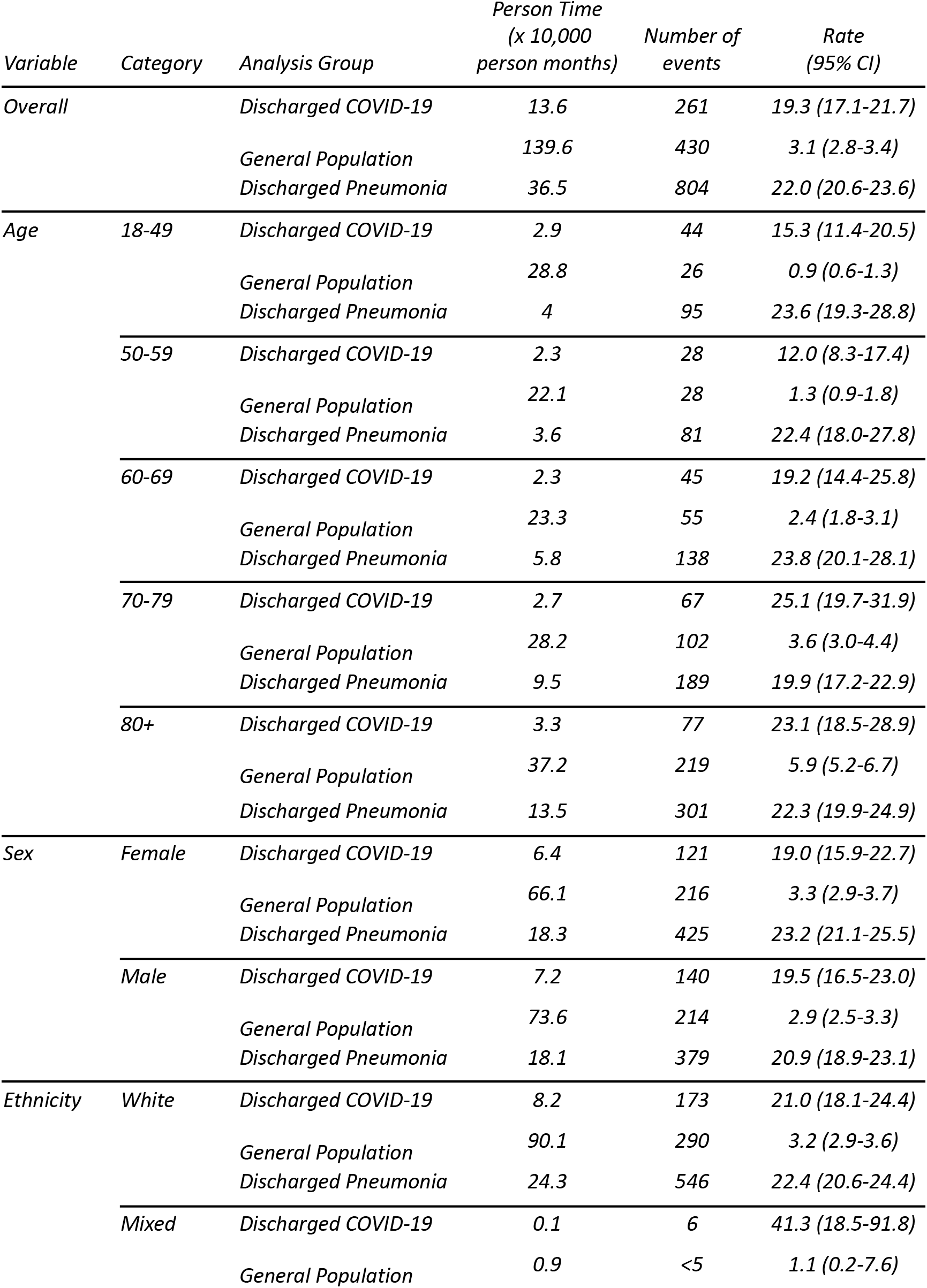
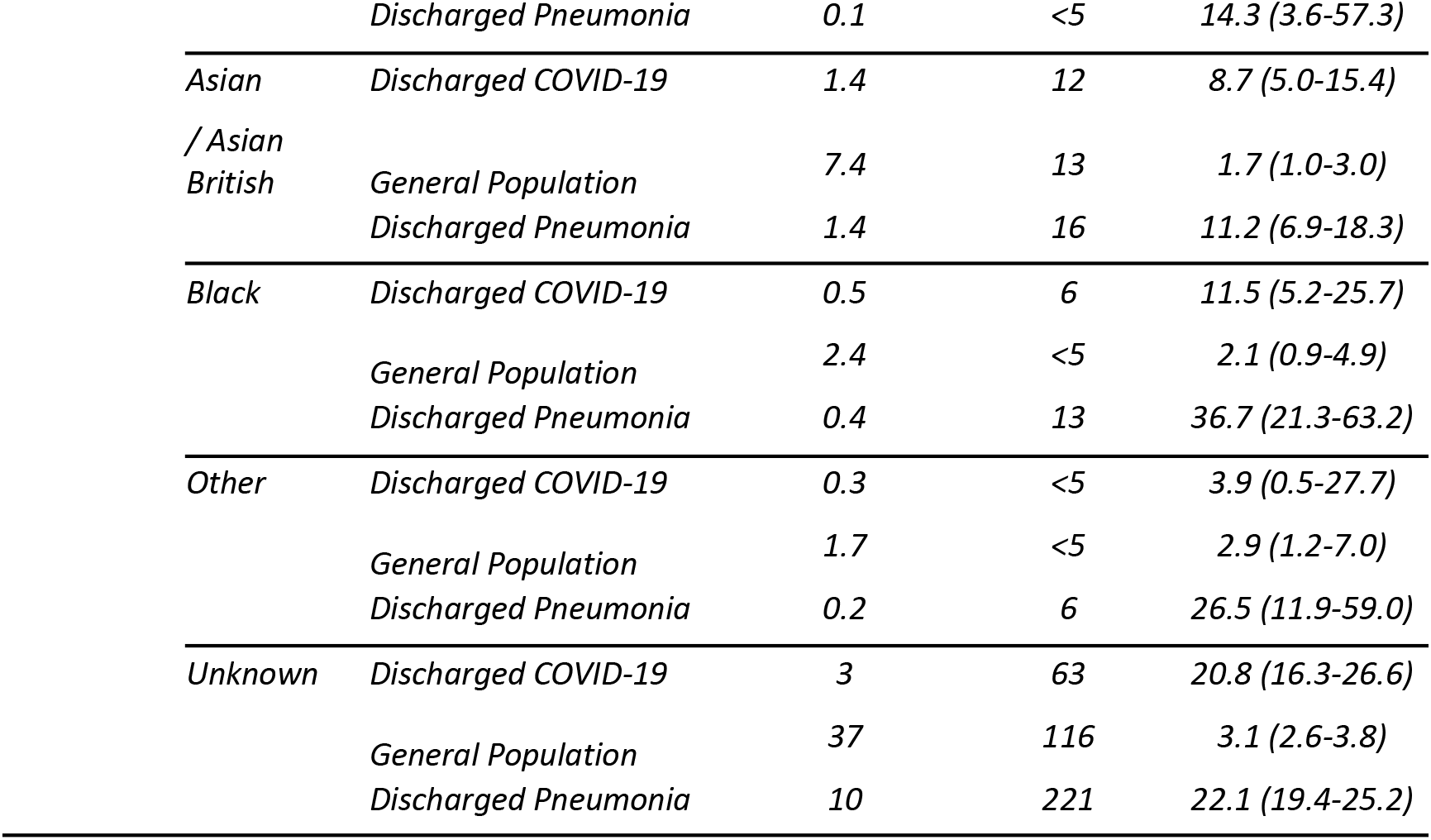
Rates of DVT amongst patients discharged with COVID-19 in 2020, all patients discharged with non-COVID pneumonia in 2019 and a matched (age, sex and region) general population comparator group in 2019

**Table A7:**
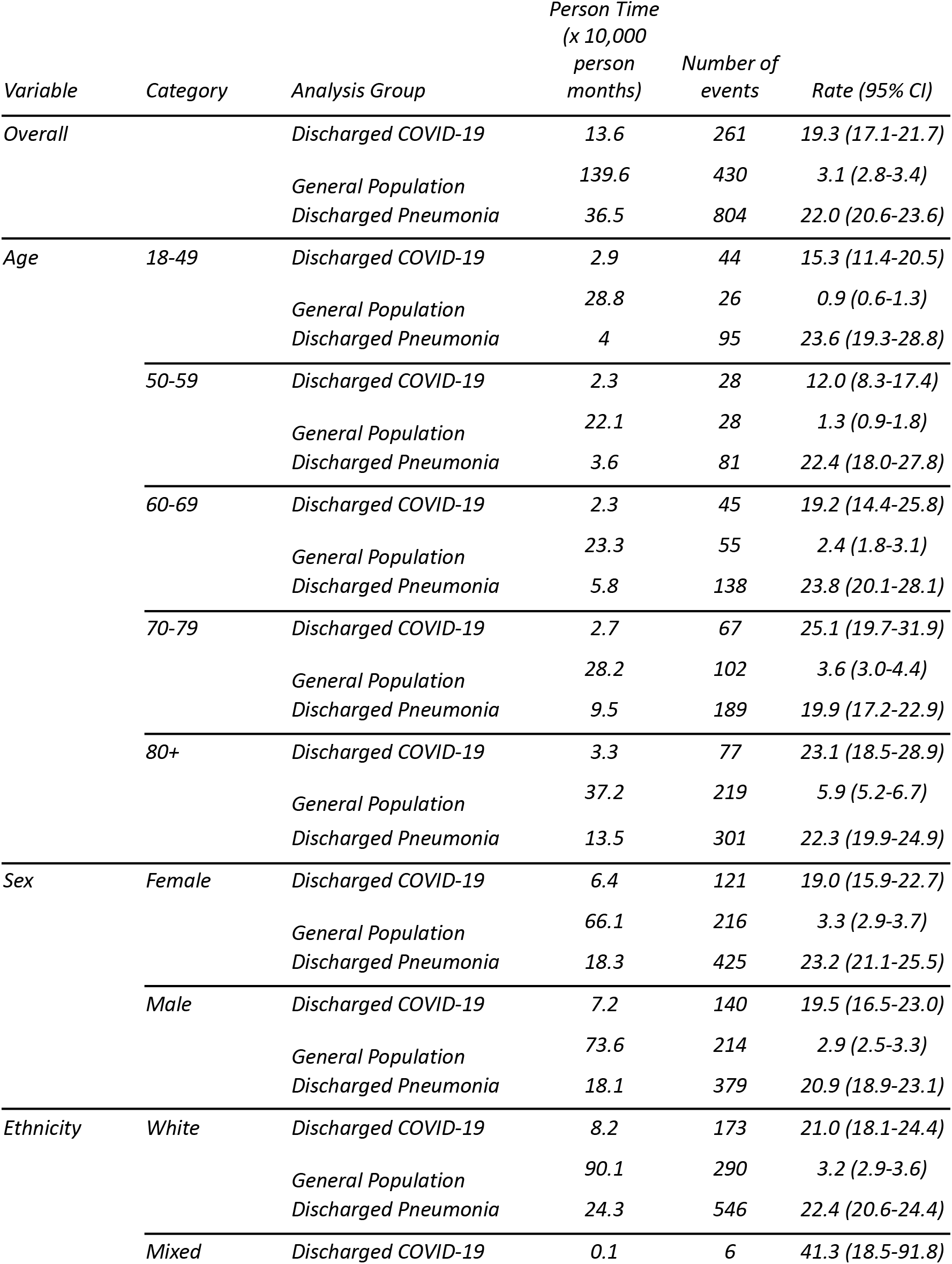
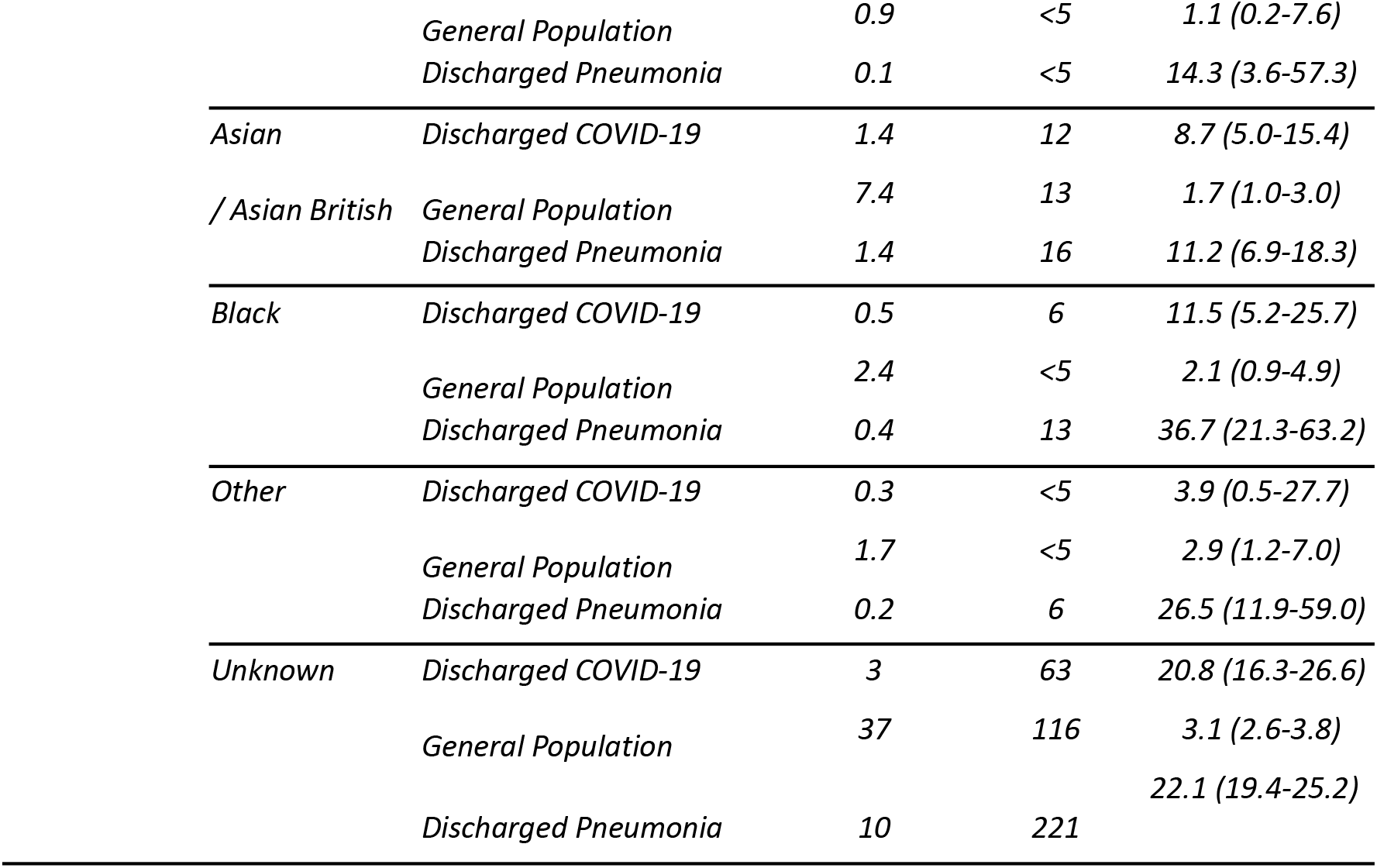
Rates of PE amongst patients discharged with COVID-19 in 2020, all patients discharged with non-COVID pneumonia in 2019 and a matched (age, sex and region) general population comparator group in 2019

| *Variable* | *Category* | *Analysis Group* | *Person Time*<br>*(x 10,000*<br>*person*<br>*months)* | *Number of*<br>*events* | *Rate (95% CI)* |
| --- | --- | --- | --- | --- | --- |
| *Overall* |  | *Discharged COVID-19* | 13.6 | 261 | 19.3 (17.1-21.7) |
|  |  | *General Population* | 139.6 | 430 | 3.1 (2.8-3.4) |
|  |  | *Discharged Pneumonia* | 36.5 | 804 | 22.0 (20.6-23.6) |
| *Age* | *18-49* | *Discharged COVID-19* | 2.9 | 44 | 15.3 (11.4-20.5) |
|  |  | *General Population* | 28.8 | 26 | 0.9 (0.6-1.3) |
|  |  | *Discharged Pneumonia* | 4 | 95 | 23.6 (19.3-28.8) |
|  | *50-59* | *Discharged COVID-19* | 2.3 | 28 | 12.0 (8.3-17.4) |
|  |  | *General Population* | 22.1 | 28 | 1.3 (0.9-1.8) |
|  |  | *Discharged Pneumonia* | 3.6 | 81 | 22.4 (18.0-27.8) |
|  | *60-69* | *Discharged COVID-19* | 2.3 | 45 | 19.2 (14.4-25.8) |
|  |  | *General Population* | 23.3 | 55 | 2.4 (1.8-3.1) |
|  |  | *Discharged Pneumonia* | 5.8 | 138 | 23.8 (20.1-28.1) |
|  | *70-79* | *Discharged COVID-19* | 2.7 | 67 | 25.1 (19.7-31.9) |
|  |  | *General Population* | 28.2 | 102 | 3.6 (3.0-4.4) |
|  |  | *Discharged Pneumonia* | 9.5 | 189 | 19.9 (17.2-22.9) |
|  | *80+* | *Discharged COVID-19* | 3.3 | 77 | 23.1 (18.5-28.9) |
|  |  | *General Population* | 37.2 | 219 | 5.9 (5.2-6.7) |
|  |  | *Discharged Pneumonia* | 13.5 | 301 | 22.3 (19.9-24.9) |
| *Sex* | *Female* | *Discharged COVID-19* | 6.4 | 121 | 19.0 (15.9-22.7) |
|  |  | *General Population* | 66.1 | 216 | 3.3 (2.9-3.7) |
|  |  | *Discharged Pneumonia* | 18.3 | 425 | 23.2 (21.1-25.5) |
|  | *Male* | *Discharged COVID-19* | 7.2 | 140 | 19.5 (16.5-23.0) |
|  |  | *General Population* | 73.6 | 214 | 2.9 (2.5-3.3) |
|  |  | *Discharged Pneumonia* | 18.1 | 379 | 20.9 (18.9-23.1) |
| *Ethnicity* | *White* | *Discharged COVID-19* | 8.2 | 173 | 21.0 (18.1-24.4) |
|  |  | *General Population* | 90.1 | 290 | 3.2 (2.9-3.6) |
|  |  | *Discharged Pneumonia* | 24.3 | 546 | 22.4 (20.6-24.4) |
|  | *Mixed* | *Discharged COVID-19* | 0.1 | 6 | 41.3 (18.5-91.8) |

|  |  |  |  |  |
| --- | --- | --- | --- | --- |
|  | *General Population* | 0.9 | <5 | 1.1 (0.2-7.6) |
|  | *Discharged Pneumonia* | 0.1 | <5 | 14.3 (3.6-57.3) |
| *Asian* | *Discharged COVID-19* | 1.4 | 12 | 8.7 (5.0-15.4) |
| */ Asian British* | *General Population* | 7.4 | 13 | 1.7 (1.0-3.0) |
|  | *Discharged Pneumonia* | 1.4 | 16 | 11.2 (6.9-18.3) |
| *Black* | *Discharged COVID-19* | 0.5 | 6 | 11.5 (5.2-25.7) |
|  | *General Population* | 2.4 | <5 | 2.1 (0.9-4.9) |
|  | *Discharged Pneumonia* | 0.4 | 13 | 36.7 (21.3-63.2) |
| *Other* | *Discharged COVID-19* | 0.3 | <5 | 3.9 (0.5-27.7) |
|  | *General Population* | 1.7 | <5 | 2.9 (1.2-7.0) |
|  | *Discharged Pneumonia* | 0.2 | 6 | 26.5 (11.9-59.0) |
| *Unknown* | *Discharged COVID-19* | 3 | 63 | 20.8 (16.3-26.6) |
|  | *General Population* | 37 | 116 | 3.1 (2.6-3.8) |
|  | *Discharged Pneumonia* | 10 | 221 | 22.1 (19.4-25.2) |

**Table A8:**
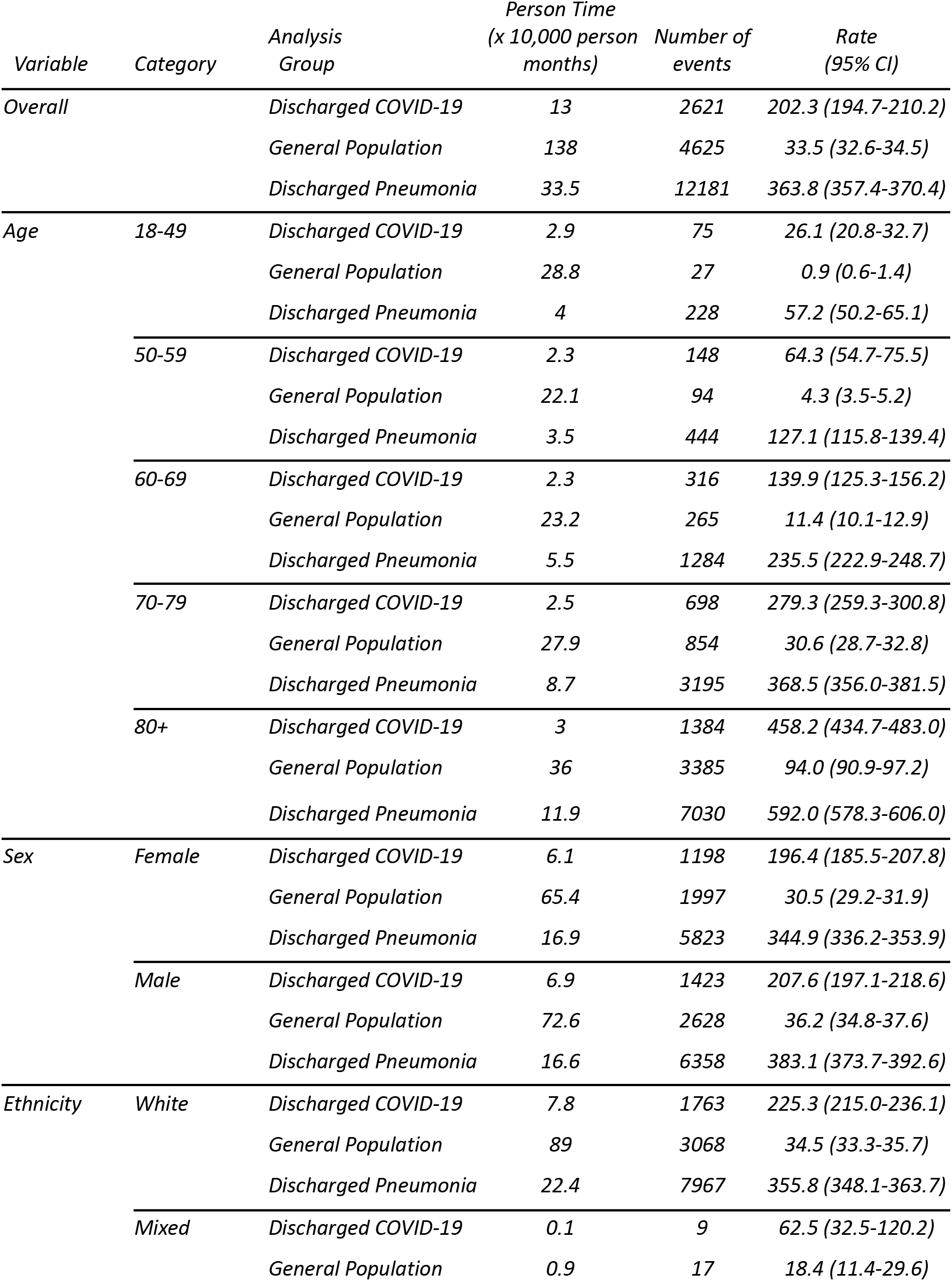
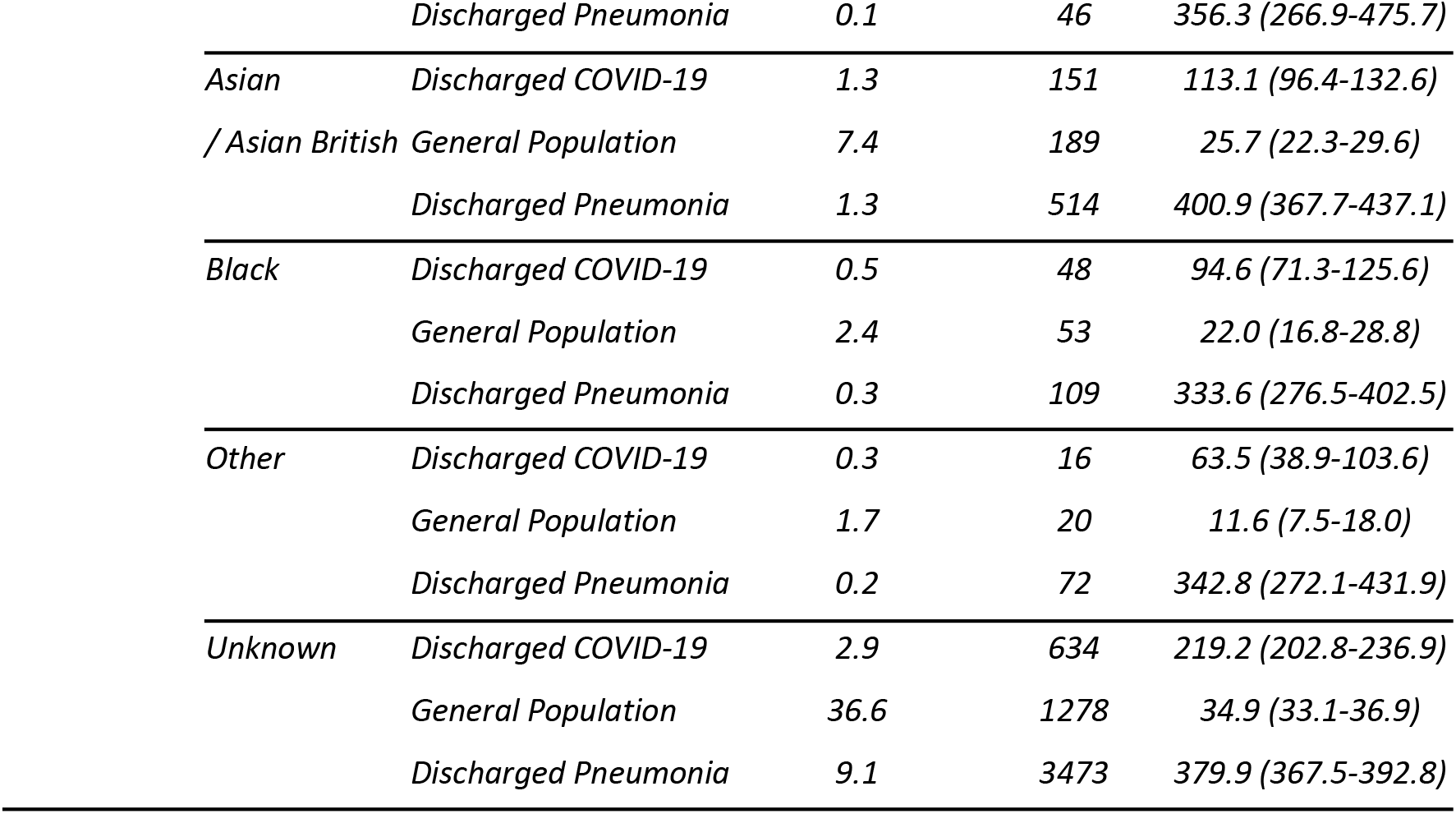
Rates of heart failure amongst patients discharged with COVID-19 in 2020, all patients discharged with non-COVID pneumonia in 2019 and a matched (age, sex and region) general population comparator group in 2019

| *Variable* | *Category* | *Analysis Group* | *Person Time*<br>*(x 10,000 person*<br>*months)* | *Number of*<br>*events* | *Rate*<br>*(95% CI)* |
| --- | --- | --- | --- | --- | --- |
| *Overall* |  | *Discharged COVID-19* | 13 | 2621 | 202.3 (194.7-210.2) |
|  |  | *General Population* | 138 | 4625 | 33.5 (32.6-34.5) |
|  |  | *Discharged Pneumonia* | 33.5 | 12181 | 363.8 (357.4-370.4) |
| *Age* | *18-49* | *Discharged COVID-19* | 2.9 | 75 | 26.1 (20.8-32.7) |
|  |  | *General Population* | 28.8 | 27 | 0.9 (0.6-1.4) |
|  |  | *Discharged Pneumonia* | 4 | 228 | 57.2 (50.2-65.1) |
|  | *50-59* | *Discharged COVID-19* | 2.3 | 148 | 64.3 (54.7-75.5) |
|  |  | *General Population* | 22.1 | 94 | 4.3 (3.5-5.2) |
|  |  | *Discharged Pneumonia* | 3.5 | 444 | 127.1 (115.8-139.4) |
|  | *60-69* | *Discharged COVID-19* | 2.3 | 316 | 139.9 (125.3-156.2) |
|  |  | *General Population* | 23.2 | 265 | 11.4 (10.1-12.9) |
|  |  | *Discharged Pneumonia* | 5.5 | 1284 | 235.5 (222.9-248.7) |
|  | *70-79* | *Discharged COVID-19* | 2.5 | 698 | 279.3 (259.3-300.8) |
|  |  | *General Population* | 27.9 | 854 | 30.6 (28.7-32.8) |
|  |  | *Discharged Pneumonia* | 8.7 | 3195 | 368.5 (356.0-381.5) |
|  | *80+* | *Discharged COVID-19* | 3 | 1384 | 458.2 (434.7-483.0) |
|  |  | *General Population* | 36 | 3385 | 94.0 (90.9-97.2) |
|  |  | *Discharged Pneumonia* | 11.9 | 7030 | 592.0 (578.3-606.0) |
| *Sex* | *Female* | *Discharged COVID-19* | 6.1 | 1198 | 196.4 (185.5-207.8) |
|  |  | *General Population* | 65.4 | 1997 | 30.5 (29.2-31.9) |
|  |  | *Discharged Pneumonia* | 16.9 | 5823 | 344.9 (336.2-353.9) |
|  | *Male* | *Discharged COVID-19* | 6.9 | 1423 | 207.6 (197.1-218.6) |
|  |  | *General Population* | 72.6 | 2628 | 36.2 (34.8-37.6) |
|  |  | *Discharged Pneumonia* | 16.6 | 6358 | 383.1 (373.7-392.6) |
| *Ethnicity* | *White* | *Discharged COVID-19* | 7.8 | 1763 | 225.3 (215.0-236.1) |
|  |  | *General Population* | 89 | 3068 | 34.5 (33.3-35.7) |
|  |  | *Discharged Pneumonia* | 22.4 | 7967 | 355.8 (348.1-363.7) |
|  | *Mixed* | *Discharged COVID-19* | 0.1 | 9 | 62.5 (32.5-120.2) |
|  |  | *General Population* | 0.9 | 17 | 18.4 (11.4-29.6) |

|  |  |  |  |  |
| --- | --- | --- | --- | --- |
|  | *Discharged Pneumonia* | *0.1* | *46* | *356.3 (266.9-475.7)* |
| *Asian* | *Discharged COVID-19* | *1.3* | *151* | *113.1 (96.4-132.6)* |
| */ Asian British* | *General Population* | *7.4* | *189* | *25.7 (22.3-29.6)* |
|  | *Discharged Pneumonia* | *1.3* | *514* | *400.9 (367.7-437.1)* |
| *Black* | *Discharged COVID-19* | *0.5* | *48* | *94.6 (71.3-125.6)* |
|  | *General Population* | *2.4* | *53* | *22.0 (16.8-28.8)* |
|  | *Discharged Pneumonia* | *0.3* | *109* | *333.6 (276.5-402.5)* |
| *Other* | *Discharged COVID-19* | *0.3* | *16* | *63.5 (38.9-103.6)* |
|  | *General Population* | *1.7* | *20* | *11.6 (7.5-18.0)* |
|  | *Discharged Pneumonia* | *0.2* | *72* | *342.8 (272.1-431.9)* |
| *Unknown* | *Discharged COVID-19* | *2.9* | *634* | *219.2 (202.8-236.9)* |
|  | *General Population* | *36.6* | *1278* | *34.9 (33.1-36.9)* |
|  | *Discharged Pneumonia* | *9.1* | *3473* | *379.9 (367.5-392.8)* |

**Table A9:**
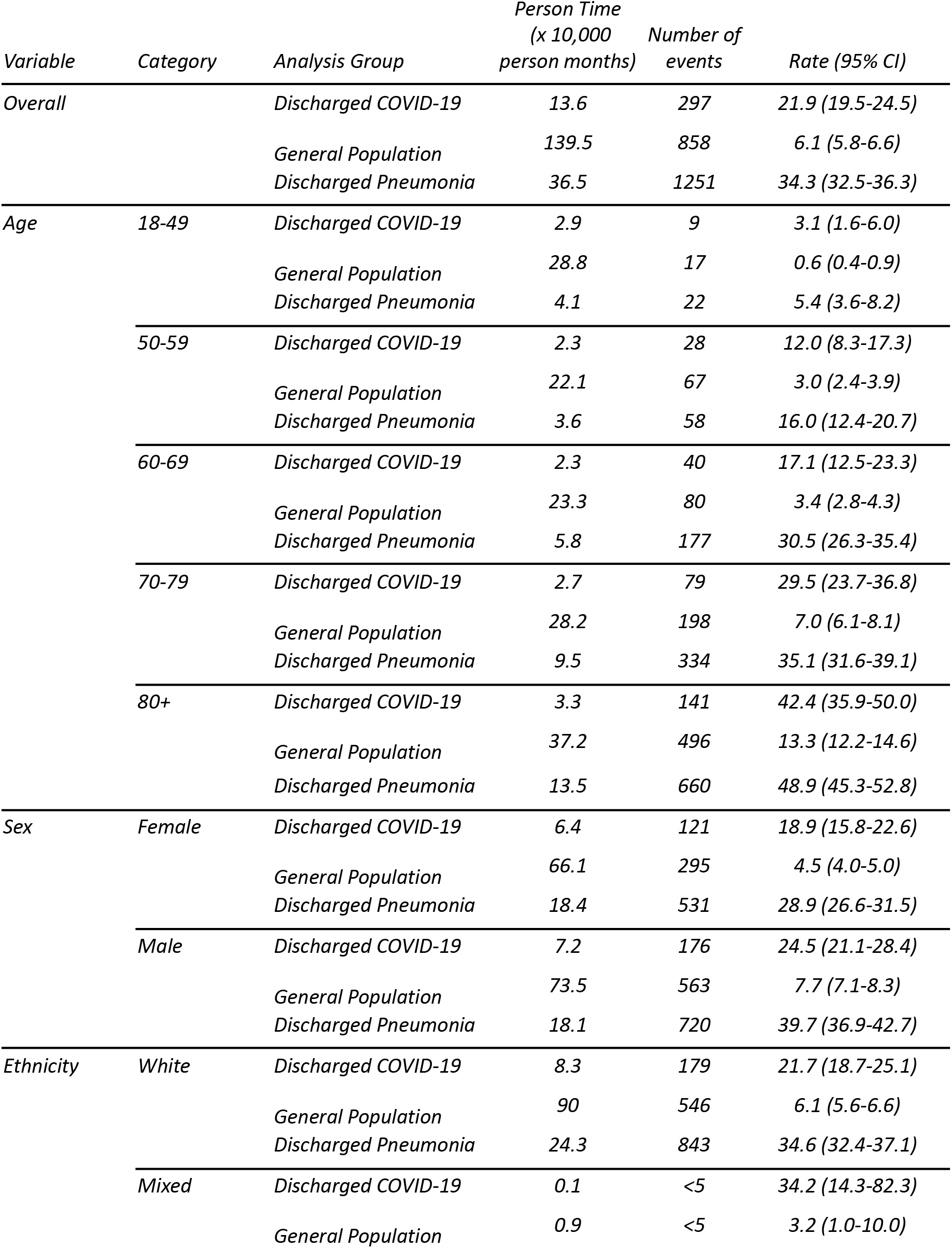
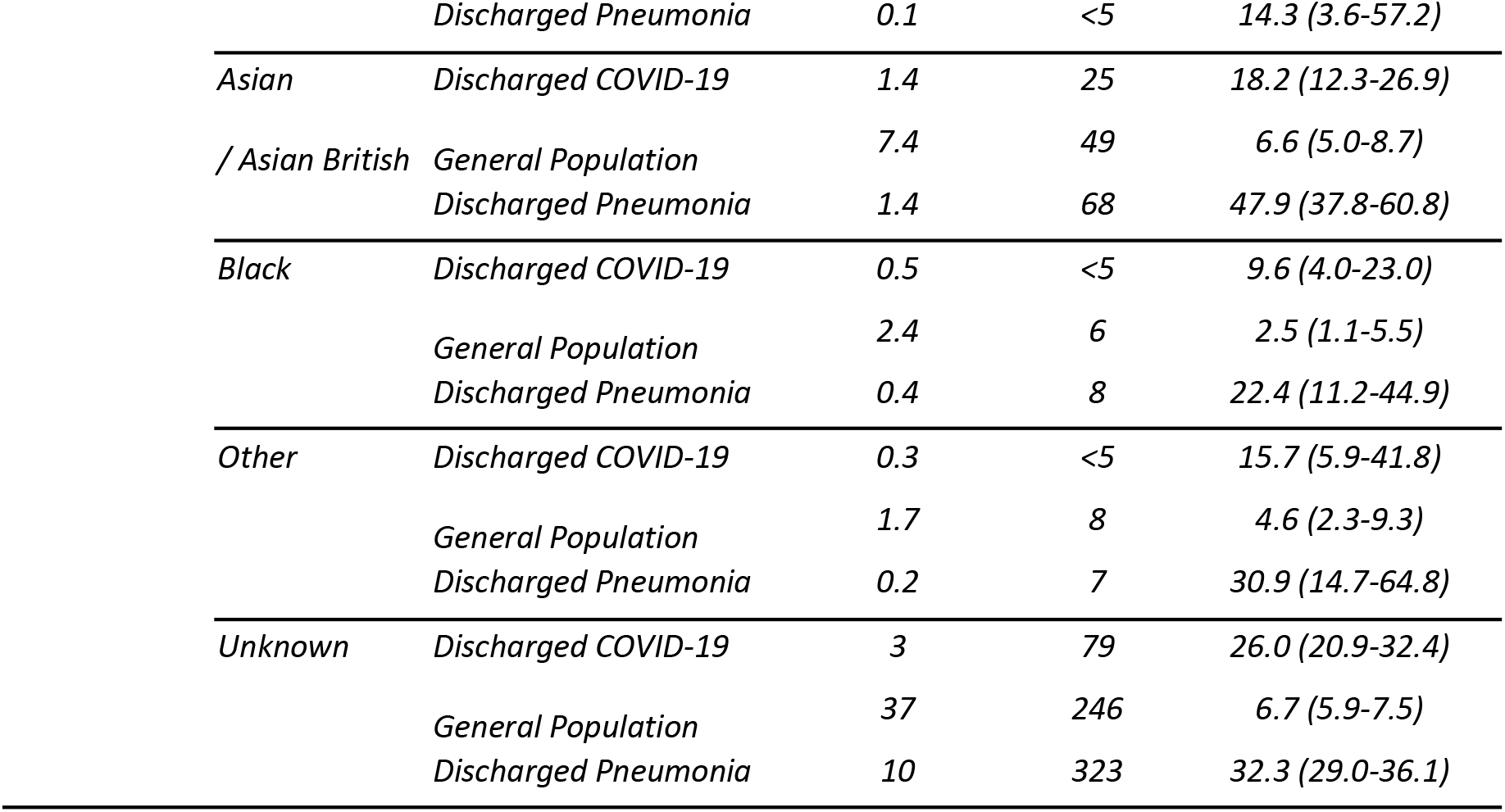
Rates of MI amongst patients discharged with COVID-19 in 2020, all patients discharged with non-COVID pneumonia in 2019 and a matched (age, sex and region) general population comparator group in 2019

| *Variable* | *Category* | *Analysis Group* | *Person Time*<br>*(x 10,000*<br>*person months)* | *Number of*<br>*events* | *Rate (95% CI)* |
| --- | --- | --- | --- | --- | --- |
| *Overall* |  | *Discharged COVID-19* | 13.6 | 297 | 21.9 (19.5-24.5) |
|  |  | *General Population* | 139.5 | 858 | 6.1 (5.8-6.6) |
|  |  | *Discharged Pneumonia* | 36.5 | 1251 | 34.3 (32.5-36.3) |
| *Age* | *18-49* | *Discharged COVID-19* | 2.9 | 9 | 3.1 (1.6-6.0) |
|  |  | *General Population* | 28.8 | 17 | 0.6 (0.4-0.9) |
|  |  | *Discharged Pneumonia* | 4.1 | 22 | 5.4 (3.6-8.2) |
|  | *50-59* | *Discharged COVID-19* | 2.3 | 28 | 12.0 (8.3-17.3) |
|  |  | *General Population* | 22.1 | 67 | 3.0 (2.4-3.9) |
|  |  | *Discharged Pneumonia* | 3.6 | 58 | 16.0 (12.4-20.7) |
|  | *60-69* | *Discharged COVID-19* | 2.3 | 40 | 17.1 (12.5-23.3) |
|  |  | *General Population* | 23.3 | 80 | 3.4 (2.8-4.3) |
|  |  | *Discharged Pneumonia* | 5.8 | 177 | 30.5 (26.3-35.4) |
|  | *70-79* | *Discharged COVID-19* | 2.7 | 79 | 29.5 (23.7-36.8) |
|  |  | *General Population* | 28.2 | 198 | 7.0 (6.1-8.1) |
|  |  | *Discharged Pneumonia* | 9.5 | 334 | 35.1 (31.6-39.1) |
|  | *80+* | *Discharged COVID-19* | 3.3 | 141 | 42.4 (35.9-50.0) |
|  |  | *General Population* | 37.2 | 496 | 13.3 (12.2-14.6) |
|  |  | *Discharged Pneumonia* | 13.5 | 660 | 48.9 (45.3-52.8) |
| *Sex* | *Female* | *Discharged COVID-19* | 6.4 | 121 | 18.9 (15.8-22.6) |
|  |  | *General Population* | 66.1 | 295 | 4.5 (4.0-5.0) |
|  |  | *Discharged Pneumonia* | 18.4 | 531 | 28.9 (26.6-31.5) |
|  | *Male* | *Discharged COVID-19* | 7.2 | 176 | 24.5 (21.1-28.4) |
|  |  | *General Population* | 73.5 | 563 | 7.7 (7.1-8.3) |
|  |  | *Discharged Pneumonia* | 18.1 | 720 | 39.7 (36.9-42.7) |
| *Ethnicity* | *White* | *Discharged COVID-19* | 8.3 | 179 | 21.7 (18.7-25.1) |
|  |  | *General Population* | 90 | 546 | 6.1 (5.6-6.6) |
|  |  | *Discharged Pneumonia* | 24.3 | 843 | 34.6 (32.4-37.1) |
|  | *Mixed* | *Discharged COVID-19* | 0.1 | <5 | 34.2 (14.3-82.3) |
|  |  | *General Population* | 0.9 | <5 | 3.2 (1.0-10.0) |

|  |  |  |  |  |
| --- | --- | --- | --- | --- |
|  | *Discharged Pneumonia* | *0.1* | *<5* | *14.3 (3.6-57.2)* |
| *Asian* | *Discharged COVID-19* | *1.4* | *25* | *18.2 (12.3-26.9)* |
| */ Asian British* | *General Population* | *7.4* | *49* | *6.6 (5.0-8.7)* |
|  | *Discharged Pneumonia* | *1.4* | *68* | *47.9 (37.8-60.8)* |
| *Black* | *Discharged COVID-19* | *0.5* | *<5* | *9.6 (4.0-23.0)* |
|  | *General Population* | *2.4* | *6* | *2.5 (1.1-5.5)* |
|  | *Discharged Pneumonia* | *0.4* | *8* | *22.4 (11.2-44.9)* |
| *Other* | *Discharged COVID-19* | *0.3* | *<5* | *15.7 (5.9-41.8)* |
|  | *General Population* | *1.7* | *8* | *4.6 (2.3-9.3)* |
|  | *Discharged Pneumonia* | *0.2* | *7* | *30.9 (14.7-64.8)* |
| *Unknown* | *Discharged COVID-19* | *3* | *79* | *26.0 (20.9-32.4)* |
|  | *General Population* | *37* | *246* | *6.7 (5.9-7.5)* |
|  | *Discharged Pneumonia* | *10* | *323* | *32.3 (29.0-36.1)* |

**Table A10:**
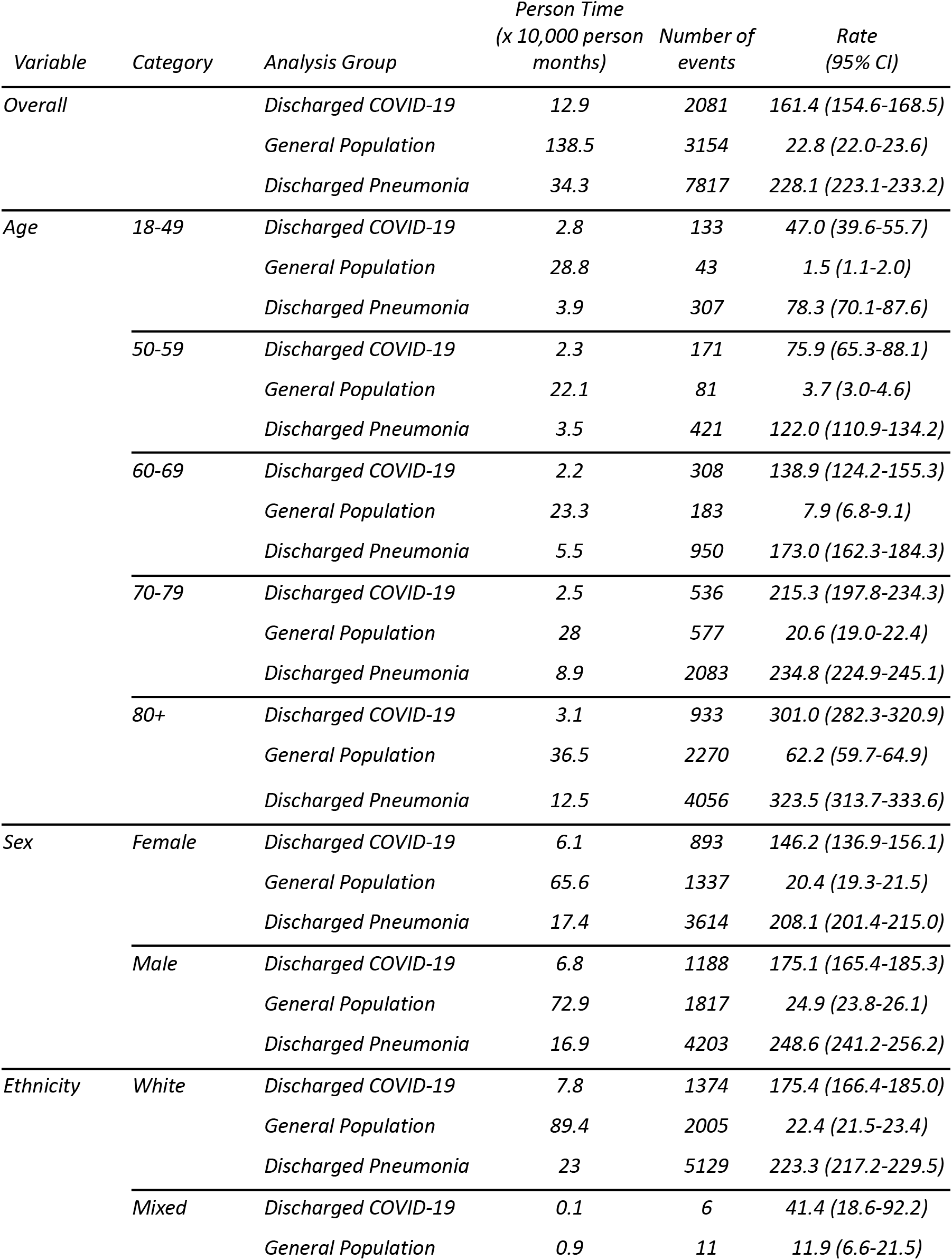
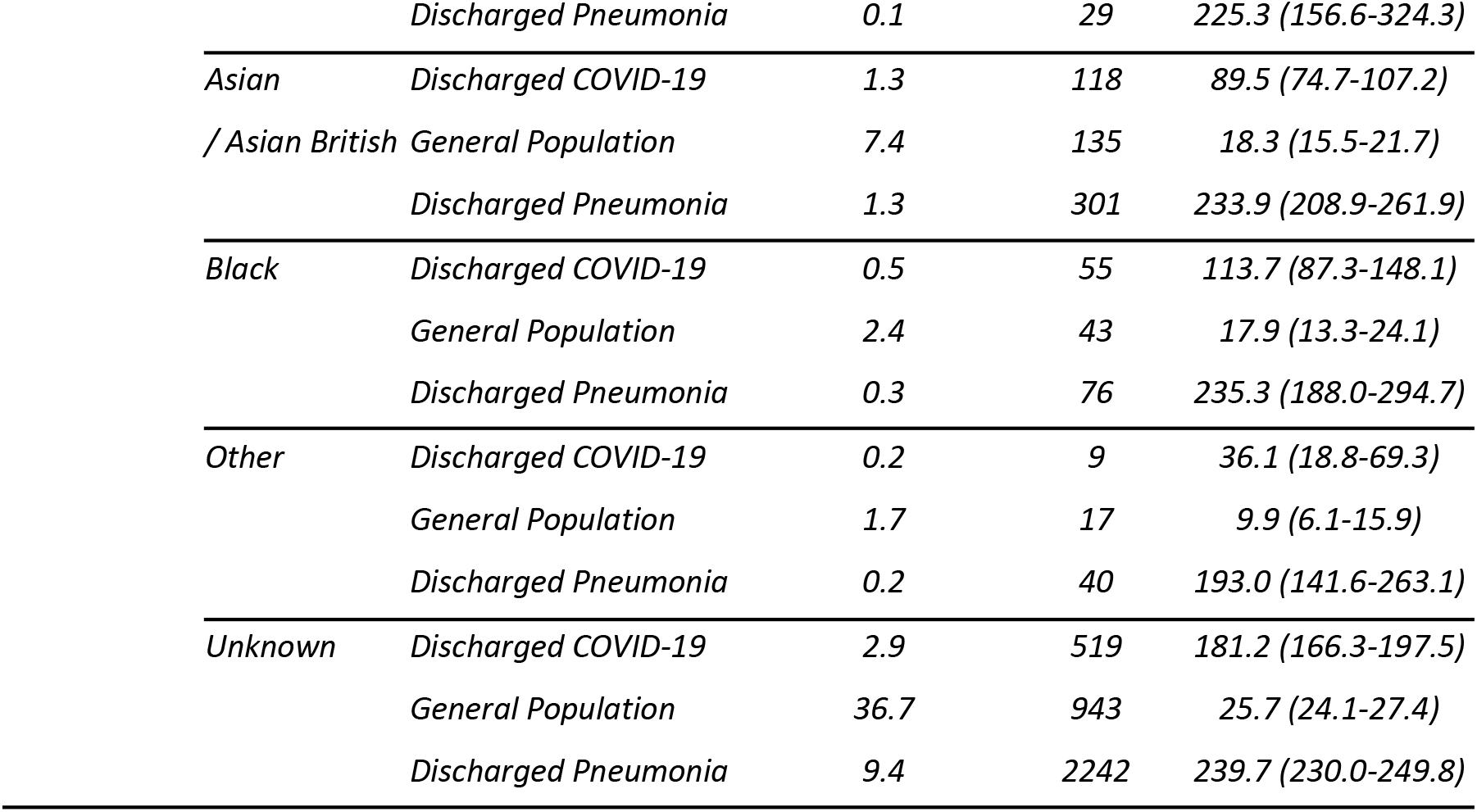
Rates of AKI amongst patients discharged with COVID-19 in 2020, all patients discharged with non-COVID pneumonia in 2019 and a matched (age, sex and region) general population comparator group in 2019

| *Variable* | *Category* | *Analysis Group* | *Person Time*<br>*(x 10,000 person*<br>*months)* | *Number of*<br>*events* | *Rate*<br>*(95% CI)* |
| --- | --- | --- | --- | --- | --- |
| *Overall* |  | *Discharged COVID-19* | 12.9 | 2081 | 161.4 (154.6-168.5) |
|  |  | *General Population* | 138.5 | 3154 | 22.8 (22.0-23.6) |
|  |  | *Discharged Pneumonia* | 34.3 | 7817 | 228.1 (223.1-233.2) |
| *Age* | *18-49* | *Discharged COVID-19* | 2.8 | 133 | 47.0 (39.6-55.7) |
|  |  | *General Population* | 28.8 | 43 | 1.5 (1.1-2.0) |
|  |  | *Discharged Pneumonia* | 3.9 | 307 | 78.3 (70.1-87.6) |
|  | *50-59* | *Discharged COVID-19* | 2.3 | 171 | 75.9 (65.3-88.1) |
|  |  | *General Population* | 22.1 | 81 | 3.7 (3.0-4.6) |
|  |  | *Discharged Pneumonia* | 3.5 | 421 | 122.0 (110.9-134.2) |
|  | *60-69* | *Discharged COVID-19* | 2.2 | 308 | 138.9 (124.2-155.3) |
|  |  | *General Population* | 23.3 | 183 | 7.9 (6.8-9.1) |
|  |  | *Discharged Pneumonia* | 5.5 | 950 | 173.0 (162.3-184.3) |
|  | *70-79* | *Discharged COVID-19* | 2.5 | 536 | 215.3 (197.8-234.3) |
|  |  | *General Population* | 28 | 577 | 20.6 (19.0-22.4) |
|  |  | *Discharged Pneumonia* | 8.9 | 2083 | 234.8 (224.9-245.1) |
|  | *80+* | *Discharged COVID-19* | 3.1 | 933 | 301.0 (282.3-320.9) |
|  |  | *General Population* | 36.5 | 2270 | 62.2 (59.7-64.9) |
|  |  | *Discharged Pneumonia* | 12.5 | 4056 | 323.5 (313.7-333.6) |
| *Sex* | *Female* | *Discharged COVID-19* | 6.1 | 893 | 146.2 (136.9-156.1) |
|  |  | *General Population* | 65.6 | 1337 | 20.4 (19.3-21.5) |
|  |  | *Discharged Pneumonia* | 17.4 | 3614 | 208.1 (201.4-215.0) |
|  | *Male* | *Discharged COVID-19* | 6.8 | 1188 | 175.1 (165.4-185.3) |
|  |  | *General Population* | 72.9 | 1817 | 24.9 (23.8-26.1) |
|  |  | *Discharged Pneumonia* | 16.9 | 4203 | 248.6 (241.2-256.2) |
| *Ethnicity* | *White* | *Discharged COVID-19* | 7.8 | 1374 | 175.4 (166.4-185.0) |
|  |  | *General Population* | 89.4 | 2005 | 22.4 (21.5-23.4) |
|  |  | *Discharged Pneumonia* | 23 | 5129 | 223.3 (217.2-229.5) |
|  | *Mixed* | *Discharged COVID-19* | 0.1 | 6 | 41.4 (18.6-92.2) |
|  |  | *General Population* | 0.9 | 11 | 11.9 (6.6-21.5) |

|  |  |  |  |  |
| --- | --- | --- | --- | --- |
|  | *Discharged Pneumonia* | *0.1* | *29* | *225.3 (156.6-324.3)* |
| *Asian* | *Discharged COVID-19* | *1.3* | *118* | *89.5 (74.7-107.2)* |
| */ Asian British* | *General Population* | *7.4* | *135* | *18.3 (15.5-21.7)* |
|  | *Discharged Pneumonia* | *1.3* | *301* | *233.9 (208.9-261.9)* |
| *Black* | *Discharged COVID-19* | *0.5* | *55* | *113.7 (87.3-148.1)* |
|  | *General Population* | *2.4* | *43* | *17.9 (13.3-24.1)* |
|  | *Discharged Pneumonia* | *0.3* | *76* | *235.3 (188.0-294.7)* |
| *Other* | *Discharged COVID-19* | *0.2* | *9* | *36.1 (18.8-69.3)* |
|  | *General Population* | *1.7* | *17* | *9.9 (6.1-15.9)* |
|  | *Discharged Pneumonia* | *0.2* | *40* | *193.0 (141.6-263.1)* |
| *Unknown* | *Discharged COVID-19* | *2.9* | *519* | *181.2 (166.3-197.5)* |
|  | *General Population* | *36.7* | *943* | *25.7 (24.1-27.4)* |
|  | *Discharged Pneumonia* | *9.4* | *2242* | *239.7 (230.0-249.8)* |

**Table A11:**
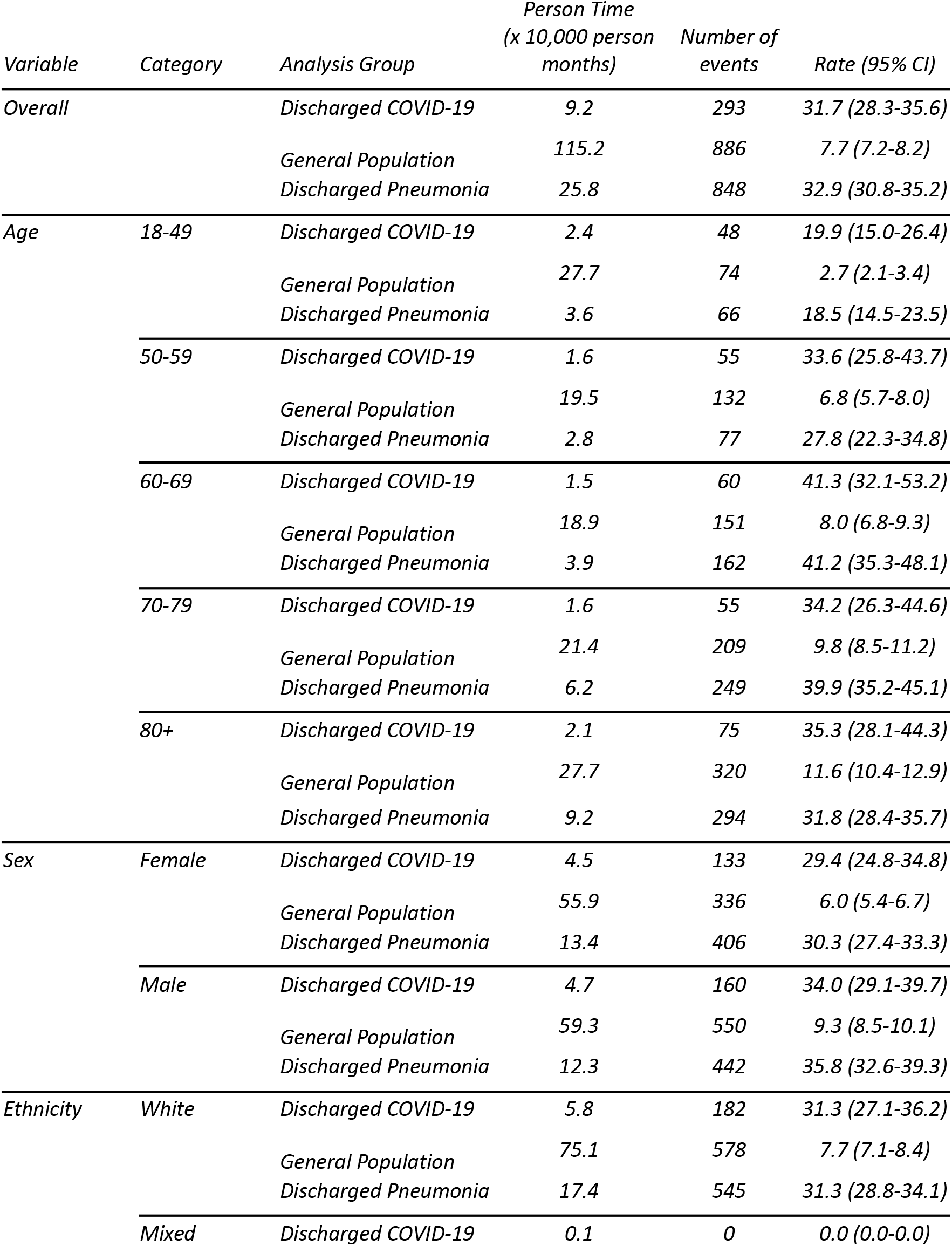
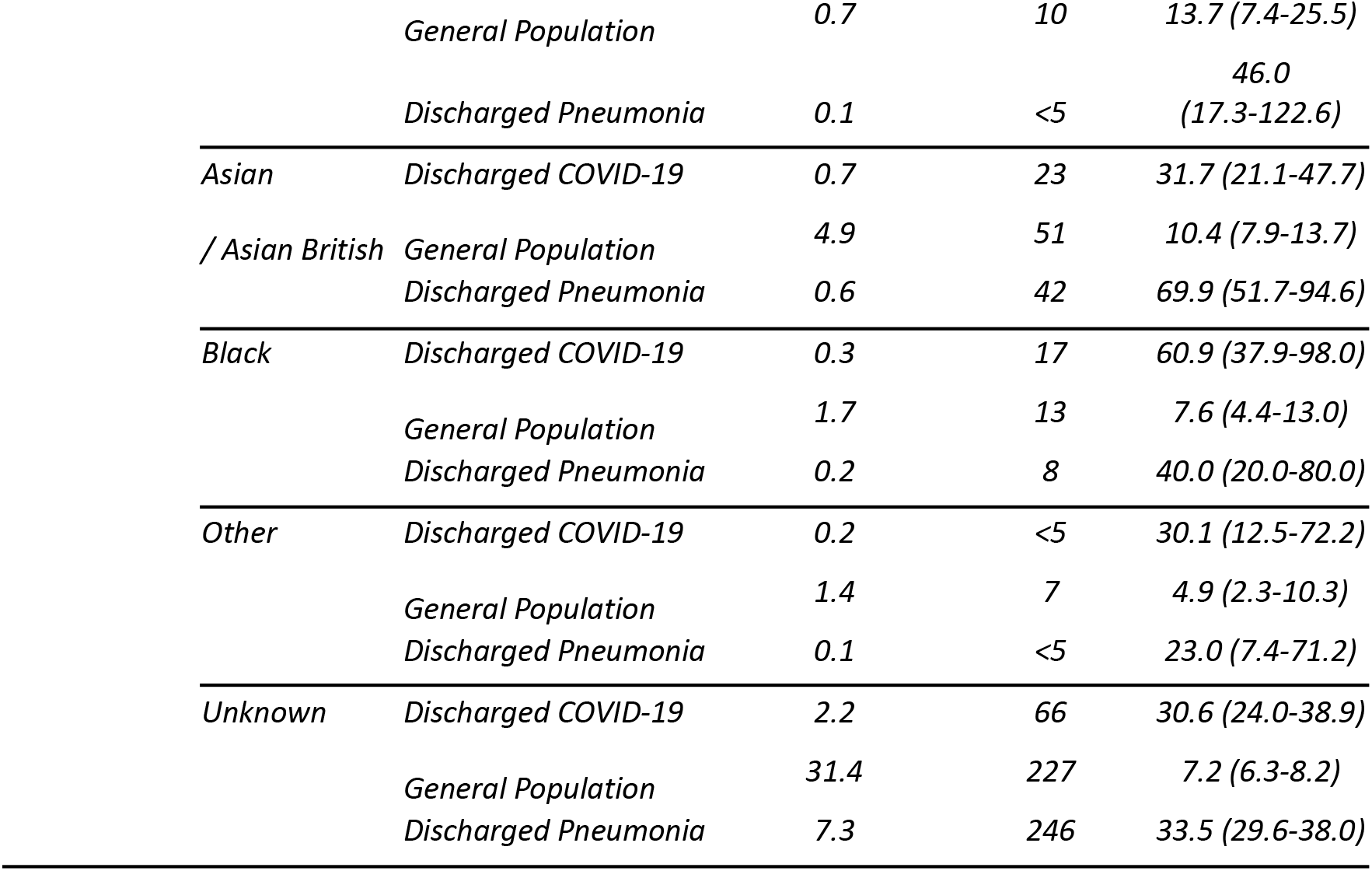
Rates of T2DM amongst patients discharged with COVID-19 in 2020, all patients discharged with non-COVID pneumonia in 2019 and a matched (age, sex and region) general population comparator group in 2019

| *Variable* | *Category* | *Analysis Group* | *Person Time*<br>*(x 10,000 person*<br>*months)* | *Number of*<br>*events* | *Rate (95% CI)* |
| --- | --- | --- | --- | --- | --- |
| *Overall* |  | *Discharged COVID-19* | 9.2 | 293 | 31.7 (28.3-35.6) |
|  |  | *General Population* | 115.2 | 886 | 7.7 (7.2-8.2) |
|  |  | *Discharged Pneumonia* | 25.8 | 848 | 32.9 (30.8-35.2) |
| *Age* | *18-49* | *Discharged COVID-19* | 2.4 | 48 | 19.9 (15.0-26.4) |
|  |  | *General Population* | 27.7 | 74 | 2.7 (2.1-3.4) |
|  |  | *Discharged Pneumonia* | 3.6 | 66 | 18.5 (14.5-23.5) |
|  | *50-59* | *Discharged COVID-19* | 1.6 | 55 | 33.6 (25.8-43.7) |
|  |  | *General Population* | 19.5 | 132 | 6.8 (5.7-8.0) |
|  |  | *Discharged Pneumonia* | 2.8 | 77 | 27.8 (22.3-34.8) |
|  | *60-69* | *Discharged COVID-19* | 1.5 | 60 | 41.3 (32.1-53.2) |
|  |  | *General Population* | 18.9 | 151 | 8.0 (6.8-9.3) |
|  |  | *Discharged Pneumonia* | 3.9 | 162 | 41.2 (35.3-48.1) |
|  | *70-79* | *Discharged COVID-19* | 1.6 | 55 | 34.2 (26.3-44.6) |
|  |  | *General Population* | 21.4 | 209 | 9.8 (8.5-11.2) |
|  |  | *Discharged Pneumonia* | 6.2 | 249 | 39.9 (35.2-45.1) |
|  | *80+* | *Discharged COVID-19* | 2.1 | 75 | 35.3 (28.1-44.3) |
|  |  | *General Population* | 27.7 | 320 | 11.6 (10.4-12.9) |
|  |  | *Discharged Pneumonia* | 9.2 | 294 | 31.8 (28.4-35.7) |
| *Sex* | *Female* | *Discharged COVID-19* | 4.5 | 133 | 29.4 (24.8-34.8) |
|  |  | *General Population* | 55.9 | 336 | 6.0 (5.4-6.7) |
|  |  | *Discharged Pneumonia* | 13.4 | 406 | 30.3 (27.4-33.3) |
|  | *Male* | *Discharged COVID-19* | 4.7 | 160 | 34.0 (29.1-39.7) |
|  |  | *General Population* | 59.3 | 550 | 9.3 (8.5-10.1) |
|  |  | *Discharged Pneumonia* | 12.3 | 442 | 35.8 (32.6-39.3) |
| *Ethnicity* | *White* | *Discharged COVID-19* | 5.8 | 182 | 31.3 (27.1-36.2) |
|  |  | *General Population* | 75.1 | 578 | 7.7 (7.1-8.4) |
|  |  | *Discharged Pneumonia* | 17.4 | 545 | 31.3 (28.8-34.1) |
|  | *Mixed* | *Discharged COVID-19* | 0.1 | 0 | 0.0 (0.0-0.0) |

|  |  |  |  |  |
| --- | --- | --- | --- | --- |
|  | *General Population* | 0.7 | 10 | 13.7 (7.4-25.5) |
|  | *Discharged Pneumonia* | 0.1 | <5 | 46.0<br>(17.3-122.6) |
| *Asian* | *Discharged COVID-19* | 0.7 | 23 | 31.7 (21.1-47.7) |
| */ Asian British* | *General Population* | 4.9 | 51 | 10.4 (7.9-13.7) |
|  | *Discharged Pneumonia* | 0.6 | 42 | 69.9 (51.7-94.6) |
| *Black* | *Discharged COVID-19* | 0.3 | 17 | 60.9 (37.9-98.0) |
|  | *General Population* | 1.7 | 13 | 7.6 (4.4-13.0) |
|  | *Discharged Pneumonia* | 0.2 | 8 | 40.0 (20.0-80.0) |
| *Other* | *Discharged COVID-19* | 0.2 | <5 | 30.1 (12.5-72.2) |
|  | *General Population* | 1.4 | 7 | 4.9 (2.3-10.3) |
|  | *Discharged Pneumonia* | 0.1 | <5 | 23.0 (7.4-71.2) |
| *Unknown* | *Discharged COVID-19* | 2.2 | 66 | 30.6 (24.0-38.9) |
|  | *General Population* | 31.4 | 227 | 7.2 (6.3-8.2) |
|  | *Discharged Pneumonia* | 7.3 | 246 | 33.5 (29.6-38.0) |

